# Lower hypothalamus subunit volumes link with long-term growth failure after preterm birth: a longitudinal case-control study

**DOI:** 10.1101/2022.01.12.22269172

**Authors:** Tobias Ruzok, Benita Schmitz-Koep, Aurore Menegaux, Robert Eves, Marcel Daamen, Henning Boecker, Esther Rieger-Fackeldey, Josef Priller, Claus Zimmer, Peter Bartmann, Dieter Wolke, Christian Sorg, Dennis M. Hedderich

## Abstract

**Background:** Preterm birth is associated with an increased risk for impaired growth. While it is known that in prematurity several somatic to environmental factors such as endocrine factors or nutrition modulate short- and long-term growth failure, the contribution of potentially impaired growth control in the brain remains elusive. We hypothesized that the structure of hypothalamic nuclei involved in growth control is altered after preterm birth, with these alterations being associated with aberrant weight development into adulthood.

**Methods:** We assessed 101 very preterm (i.e., <32 weeks of gestational age) and/or very low birth weight (i.e., <1500g; VP/VLBW) and 110 full-term born (FT) adults of the population-based Bavarian Longitudinal Study with T1-weighted MRI, deep learning-based hypothalamus subunit segmentation, and multiple body weight assessments from birth into adulthood.

**Results:** Volumes of the whole hypothalamus and hypothalamus subunits relevant for growth control were reduced in VP/VLBW adults and associated with birth variables (i.e., gestational age and intensity of neonatal treatment), body weight (i.e., weight at birth and adulthood), and growth trajectories (i.e., trajectory slopes and trajectories of those small for gestational age and with long-term catch-up growth). Concerning VP/VLBW weight trajectories, relatively larger hypothalamic volumes comparable to those of FT adults, especially in subunits including the lateral hypothalamus, were associated with favorable long-term growth trajectories.

**Conclusions:** Results demonstrate lower volumes of growth control-related hypothalamus sub-units after preterm birth that link with long-term growth failure. Data suggest postnatal development of growth-related hypothalamic nuclei in VP/VLBW individuals that corresponds with distinct body growth trajectories into adulthood.

**Funding:** This study was supported by the Deutsche Forschungsgemeinschaft (BA 6370/2-1 to C.S.), the German Federal Ministry of Education and Science (BMBF 01ER0801 to P.B. and D.W., BMBF 01ER0803 to C.S.) and the Kommission für Klinische Forschung, Technische Universität München (KKF 8700000474 to D.M.H.).

## Introduction

Preterm birth is defined as birth before 37 weeks of gestational age (GA) and is frequent with a worldwide prevalence of about 11%.^1^ Preterm birth is associated with increased risks for somatic, behavioral, and neuro-cognitive impairments, such as metabolic or cardio-vascular disorders^2, 3^ or lower IQ.^4, 5^ Concerning metabolic impairments, preterm birth elevates the risk for growth failure, with increasing risks for those born very preterm (i.e., <32 GA) and/or with very low birth weight (i.e., <1500g; VP/VLBW); postnatal to long-term growth failure is broadly defined as the failure to achieve the growth potential expected for an individual at a certain age.^6–8^ While progress in modern neonatal management overcomes postnatal differences in body height, body weight differences of VP/VLBW compared to full-term born (FT) infants are not yet compensated.^9^ Particularly, VP/VLBW infants born below the 10th percentile of weight for their gestational age (i.e., born small for gestational age, SGA) have larger hazards for postnatal growth failure and/or failed long-term catch-up growth, which, in turn, is associated with increased risks for long-term morbidity.^8, 10, 11^ Indeed, growth failure links with increased risks for neurodevelopmental impairment, dyslipidemia, impaired glucose tolerance, and diabetes mellitus-type-II.^12–14^ Moreover, growth failure represents a complex and multifactorial condition modified by maternal, genetic, fetal, environmental, nutritional, stress-related, and endocrine factors.^10, 11, 15^ While our knowledge about environmental to somatic factors is remarkable, we still do not know, however, whether altered brain mechanisms of growth control are also associated with aberrant body weight development after preterm birth.

The hypothalamus is critically involved in growth control.^16–18^ It is a highly conserved brain structure across vertebrates, surrounding the infundibular recess of the forebrain’s third ventricle and consisting of at least 13 interconnected nuclei.^19^ Although its neurogenesis starts already during gestational week 9 in humans, its development is not terminated until the third trimester of pregnancy,^20^ with subsequent development ranging from neuronal migration, axon extension, dendritic arborization and synaptogenesis to myelination and epigenetic modifications.^21^ It is known that the hypothalamus controls a variety of basic physiological-behavioral processes, from circadian rhythms to drinking, feeding, sexual, and threat behavior.^18^ In particular, several hypothalamic nuclei, including the paraventricular, infundibular, dorso-/ventromedial nucleus, and the lateral hypothalamus, are specifically involved in the control of appetite, food intake, and growth, including body weight.^16, 17^ While most evidence about hypothalamic functions stems from animal studies (e.g.,^22^), neuroimaging studies in humans have linked altered hypothalamic structure with impaired body weight control e.g., in obesity and anorexia nervosa.^23, 24^

In-vivo imaging of the hypothalamus at a nuclear level in humans is challenging due to surrounding grey matter and its small size of about 1cm³.^25, 26^ It therefore requires both, high spatial resolution imaging and optimal delineation methodology. While previous approaches on delineation relied on manual, semi-automated or automated Bayesian or multi-atlas segmentation techniques (see overview in^27^), recent improvements in deep learning-based segmentation (i.e., automated deep convolutional neural networks)^28^ enable highly reliable hypothalamus delineation, sensitive to inter-individual differences and, most importantly, sensitive to the identification of hypothalamus subsegments that can be mapped on nuclei that are relevant for body weight control (Figure 1).

**Figure 1.**
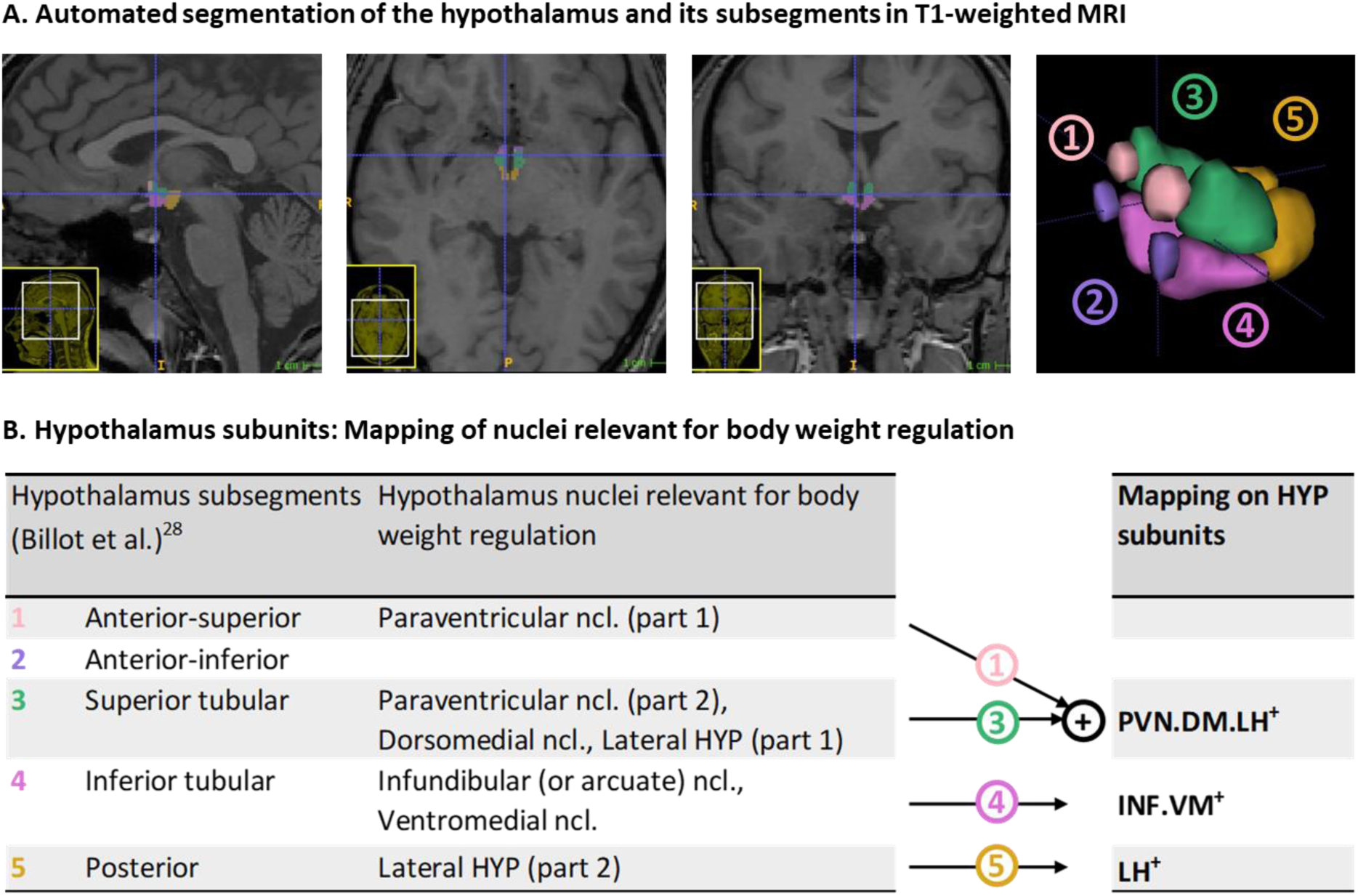
Hypothalamus segmentation and mapping on growth control relevant subunits: **(A)** Representative segmentation of the hypothalamus of a VP/VLBW adult via deep convolutional neural network algorithm in T1-weighted MRI. From left to right: sagittal, axial, coronal view, and 3D rendering of the whole hypothalamus and its subsegments. **(B)** To focus on hypothalamic nuclei involved in growth control, we mapped subsegments from the segmentation algorithm that contained nuclei relevant for growth control onto *three so-called subunits of growth control, namely PVN.DM.LH^+^, INF.VM^+^ and LH^+^*. The PVN.DM.LH^+^ subunit is the aggregate of two subsegments of the segmentation of Billot et al. (28), namely the anterior-superior and superior tubular subsegment, and these two subsegments cover the preoptic area, the paraventricular nucleus (PVN), the dorsomedial nucleus (DM), and parts of the lateral hypothalamus (LH); the latter three nuclei are involved in growth control, therefore defining the name of the subunit. INF.VM^+^ is identical to the inferior tubular subsegment, which comprises - amongst other nuclei - the growth control-related infundibular (INF) and ventromedial (VM) nuclei. LH^+^ matches the posterior subsegment in Billot et al. (2020), including mammillary bodies, and parts of the tuberomammillary nucleus, and, critically, of the growth control-related lateral hypothalamus (LH). The final anterior-inferior subsegment (suprachiasmatic nucleus and parts of the supraoptic nucleus) was excluded from further analysis because it does not contain any growth-related nuclei. All subunits stated in the analysis already consider volumes of bilateral hypothalamus. *Abbreviations:* HYP, hypothalamus; MRI, magnetic resonance imaging; VP/VLBW, very preterm and/or very low birth weight.

We assessed the role of the hypothalamus in body growth control in VP/VLBW individuals by addressing the following hypotheses: After preterm birth, (i) the structure of whole hypothalamus and hypothalamic nuclei involved in growth control is altered, and (ii) these alterations are associated (a) with aberrant weight development into adulthood, particularly in individuals (b) born SGA and (c) with long-term catch-up growth. To test these hypotheses, we assessed 101 VP/VLBW and 110 FT adults of the population-based Bavarian Longitudinal Study (BLS) with T1-weighted MRI, deep learning-based hypothalamus segmentation, hypothalamic volume as proxy for hypothalamic structure, and multiple body weight assessments, including both slope and cluster analysis of long-term body weight development trajectories, from birth to adulthood.

## Methods

### Participants

Data for this study were derived from the Bavarian Longitudinal Study, a geographically defined whole population study of neonatal at-risk VP/VLBW and FT individuals, described in detail in^29^, lastly in^30^, and in the supplement. Briefly, we used data from 101 VP/VLBW individuals and 111 FT controls who underwent MRI scan at age of 26y and life-long longitudinal body weight assessment. MRI took place at two sites, namely Department of Neuroradiology, Klinikum rechts der Isar, Technical University of Munich (n=146), and Department of Radiology, University Hospital of Bonn (n=66). For a detailed flowchart of participants through the whole study see Supplementary Fig. S1 and^30^.

### Gestation, medical impairments at birth, and body growth

We performed canonical measurements of gestational age (GA), medical treatment at birth, and body weight development, described in detail previously^30^ and in the supplement. Briefly, GA was estimated from maternal reports on the first day of the last menstrual period and from serial ultrasounds during pregnancy. To estimate medical impairments at birth, Intensity of Neonatal Treatment Index (INTI) was calculated via daily assessments of care level, respiratory support, feeding dependency, and neurological status (including mobility, muscle tone, and neurological excitability; Supplementary Table S1 for variable description). Longitudinal body weight measurements were undertaken at birth, at five and 20 months corrected for prematurity, at 56 months, 6, 8 and 26 years of chronological age, using predefined protocols.^31^ For growth analysis, we transformed body weight measurements from [g]/[kg] into z-scores relative to the exact age of each participant, typically used for the description of longitudinal changes of growth. “Small for gestational age” (SGA) versus “appropriate/large for gestational age” (AGA/LGA) was determined by birth weight (BW) z-scores being below (equivalent to z-scores < -1.282) versus above the 10th percentile, respectively.^32^ Within the SGA cohort, successful/failed endpoint catch-up growth was defined as an adult body weight z-score at age 26 above/below the 10th percentile i.e., we focused on long-term growth failure.^6^

For body growth trajectory analysis, we performed both trajectory slope and trajectory type analysis using Python version 3.7.10 and especially the “scikit-learn” package, a machine learning focused Python library. For trajectory slope analysis, linear regression of longitudinal body weight z-scores (non-interpolated) was performed to receive regressed body weight trajectories; change of body weight z-scores from birth until adulthood of regressed body weight trajectories defined delta slopes. For trajectory type analysis, we clustered VP/VLBW weight development trajectories via k-means algorithm in Python (sklearn.cluster.KMeans). The number of times the k-means algorithm was run with different centroid seeds as default initialization was N=100.

### MRI data acquisition and hypothalamus segmentation

MRI data acquisition (for details see^30^) was performed on Philips Achieva 3 T systems or Philips Ingenia 3T systems using an 8-channel SENSE head coil. Subject distribution among scanners was: Bonn Achieva 3T: 5 VP/VLBW, 11 FT; Bonn Ingenia 3T: 33 VP/VLBW, 17 FT; Munich Achieva 3T: 60 VP/VLBW, 65 FT; Munich Ingenia 3T: 3 VP/VLBW, 17 FT. Across all scanners sequence parameters were kept identical, namely high-resolution T1- weighted 3D magnetization prepared rapid acquisition gradient echo sequence with TI=1.3ms, TR=7.7ms, TE=3.9ms, flip angle=15°, 180 sagittal slices, FOV=256×256×180mm, reconstruction matrix=256×256, and reconstructed isotropic voxel size=1mm³. To account for possible confounds by scanner differences, MRI data analyses included scanner dummy variables as covariates of no interest.

T1-weighted MRI scans in Nifti-format were processed by using FreeSurfer version 7.1.1 (http://surfer.nmr.mgh.harvard.edu/), which includes a deep convolutional neural network tool of Billot et al.^28^ that enables for automated segmentation of the hypothalamus, including sub-segment parcellation (Fig. 1A). To focus on hypothalamic nuclei involved in growth control, we mapped subsegments from the segmentation algorithm that contained nuclei relevant for growth control onto three so-called subunits of growth control, namely PVN.DM.LH^+^, INF.VM^+^ and LH^+^ (see Figure 1B and legend description).

### Statistical analysis

Statistical analyses were performed using SPSS version 27 (IBM SPSS Statistics). Regarding demographical characteristics, group differences between VP/VLBW and FT cohorts were assessed using chi-square tests (sex) and two-sample t-tests (age, GA). To test whether both hypothalamic volumes or body weights are altered in prematurity, general linear models were used (dependent variable: hypothalamic volumes; fixed factor: status of prematurity; covariates: sex and additionally only for hypothalamic volume changes: scanner, total intracranial volume (TIV)). Partial correlation analysis, restricted to the VP/VLBW group and corrected for sex, scanner and TIV, was used to investigate the associations between hypothalamic volumes and variables of preterm birth or growth trajectories, respectively. To assess potential mediation effects of hypothalamic volumes regarding the association of variables of prematurity (i.e., INTI and GA, respectively) with adult body weight, a mediation analysis restricted to the VP/VLBW cohort was performed using the PROCESS toolbox (version 3.5) of SPSS. Statistical significance was set at p <0.05; all tests were two-sided. Tests were corrected for multiple comparisons for false discovery rate (FDR) according to the Benjamini-Hochberg procedure.^33^

### Data availability

The data that support the findings of this study are available from the corresponding author, upon reasonable request.

## Results

### Sample characteristics

Table 1 and Figure 2A show group demographic and growth-related variables, including longitudinal body weight measurements and their z-scores leading to growth trajectories of VP/VLBW individuals. Particularly, all longitudinal body weight measurements, including adult body weight, were significantly lower in VP/VLBW individuals compared to FT controls.

**Figure 2.**
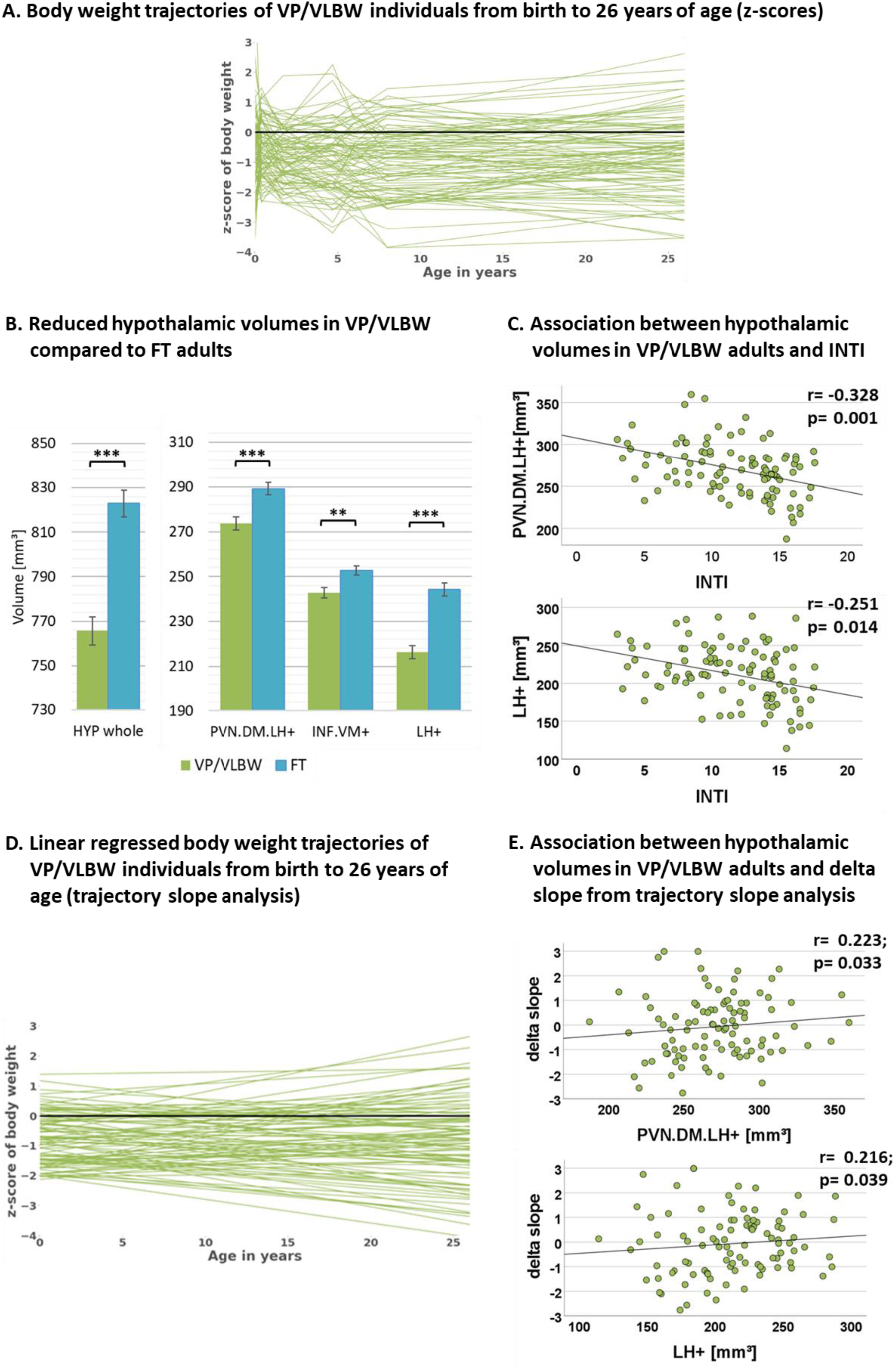
Hypothalamic volumes, prematurity, and body weight trajectories: **(A)** Individual body weight development trajectories in the VP/VLBW cohort representing z-scores of body weight at birth and age 5, 20, 56 months, 6, 8 and 26 years. **(B)** Reduced whole hypothalamus and subunit volumes in the VP/VLBW compared to the FT cohort. Marginal means of whole hypothalamus and its subunits are given in mm³ and are shown as bar plots; error bars indicate SE. Group differences were assessed using a general linear model (fixed factor: prematurity at birth, covariates of no interest: sex, scanner, TIV). Group difference significance is marked by asterisks (+: p < 0.05; *: p-FDR < 0.05; **: p-FDR < 0.01; *** p-FDR < 0.001). **(C)** Associations between hypothalamic volumes and variables of preterm birth in the VP/VLBW cohort verified by partial correlation analysis. Scatterplots show exemplary relationships between hypothalamus subunit volumes and INTI (for further results see Supplementary Table S3). **(D)** Linear regression model of individual body weight development trajectories in the VP/VLBW cohort representing body weight *slopes* via linear regression of individual z-scores of body weight at birth, age 5, 20, 56 months and 6, 8 and 26 years (trajectory slope analysis). **(E)** Association between hypothalamic volumes and delta slope in the VP/VLBW sample. Scatterplots show relationships of hypothalamus subunits with delta slope representing change of body weight z-score from birth until adulthood of linear regressed body weight trajectories. Linear regression lines and regression coefficients of partial regression analysis are added. *Abbreviations:* FDR, false discovery rate correction for multiple comparisons using the Benjamini–Hochberg method; FT, full-term; INTI, intensity of neonatal treatment; SE, standard error; VP/VLBW, very preterm and/or very low birth weight.

**Table 1:** Comparison of demographical data, TIV and body weight development variables between VP/VLBW and FT individuals.

|  |  | VP/VLBW |  |  | FT |  |  |  |
| --- | --- | --- | --- | --- | --- | --- | --- | --- |
|  | Samples | M | SD | Range | M | SD | Range | p-Value |
| Sex (male/female) | 101/110 | 58/43 |  |  | 65/45 |  |  | 0.806 |
| Age (years) | 101/110 | 26.7 | ±0.61 | 25.7 - 28.3 | 26.8 | ±0.74 | 25.5 - 28.9 | 0.165 |
| GA (weeks) | 101/110 | 30.5 | ±2.1 | 25 - 36 | 39.7 | ±1.1 | 37 - 42 | <0.001 |
| INTI | 100/ - | 11.6 | ±3.8 | 3 - 18 | n.a. | n.a. | n.a. | n.a. |
|  |  | M | SE | Range | M | SE | Range | p-Value |
| TIV (cm<sup>3</sup>) | 101/110 | 1,46 | ±11.4 | 1,44 - 1,48 | 1,53 | ±10.9 | 1,51 - 1,55 | <0.001 |
| Birth weight (g) | 97 /107 | 1,33 | ±39.3 | 1,25 - 1,40 | 3,41 | ±37.4 | 3,33 - 3,48 | <0.001 |
| Body weight, 5m (kg) | 90 /107 | 6.56 | ±0.09 | 6.40 - 6.73 | 7.38 | ±0.08 | 7.23 - 7.53 | <0.001 |
| Body weight, 20m (kg) | 97 /107 | 9.77 | ±0.26 | 9.25 - 10.3 | 11.4 | ±0.25 | 10.9 - 11.9 | <0.001 |
| Body weight, 56m (kg) | 92 /104 | 15.7 | ±0.38 | 14.9 - 16.4 | 18.2 | ±0.36 | 17.5 - 18.9 | <0.001 |
| Body weight, 6y (kg) | 84 /105 | 20.1 | ±0.32 | 19.5 - 20.8 | 21.7 | ±0.29 | 21.2 - 22.3 | <0.001 |
| Body weight, 8y (kg) | 86 /105 | 25.1 | ±0.49 | 24.1 - 26.1 | 28.0 | ±0.45 | 27.2 - 28.9 | <0.001 |
| Body weight, 26y (kg) | 97 /107 | 67.4 | ±1.4 | 64.6 - 70.2 | 77.2 | ±1.3 | 74.5 - 79.8 | <0.001 |

Statistical comparisons: Sex with $\chi^2$ statistics; Age and GA with two-sample t-tests; TIV at age 26 (covariates: sex, scanner) and all body weight related variables (covariate: sex) with general linear model.
*Abbreviations:* FT, full-term; GA, gestational age; INTI, intensity of neonatal treatment index; Samples, cohort sample size (VP/VLBW and FT); SD, standard deviation; SE, standard error; TIV, total intracranial volume; VP/VLBW, very preterm and/or very low birth weight.

### Adult hypothalamic volumes, VP/VLBW effect, and slopes of body weight trajectories

To investigate whether hypothalamic volumes, including subunits involved in body weight control, are altered after preterm birth, we applied a general linear model approach controlling for sex, scanner, and TIV. We found significantly lower volumes in whole hypothalamus, PVN.DM.LH^+^, INF.VM^+^ and LH^+^ (all p-values <0.002) in VP/VLBW versus FT adults (Figure 2B, Table 2A).

**Table 2:**
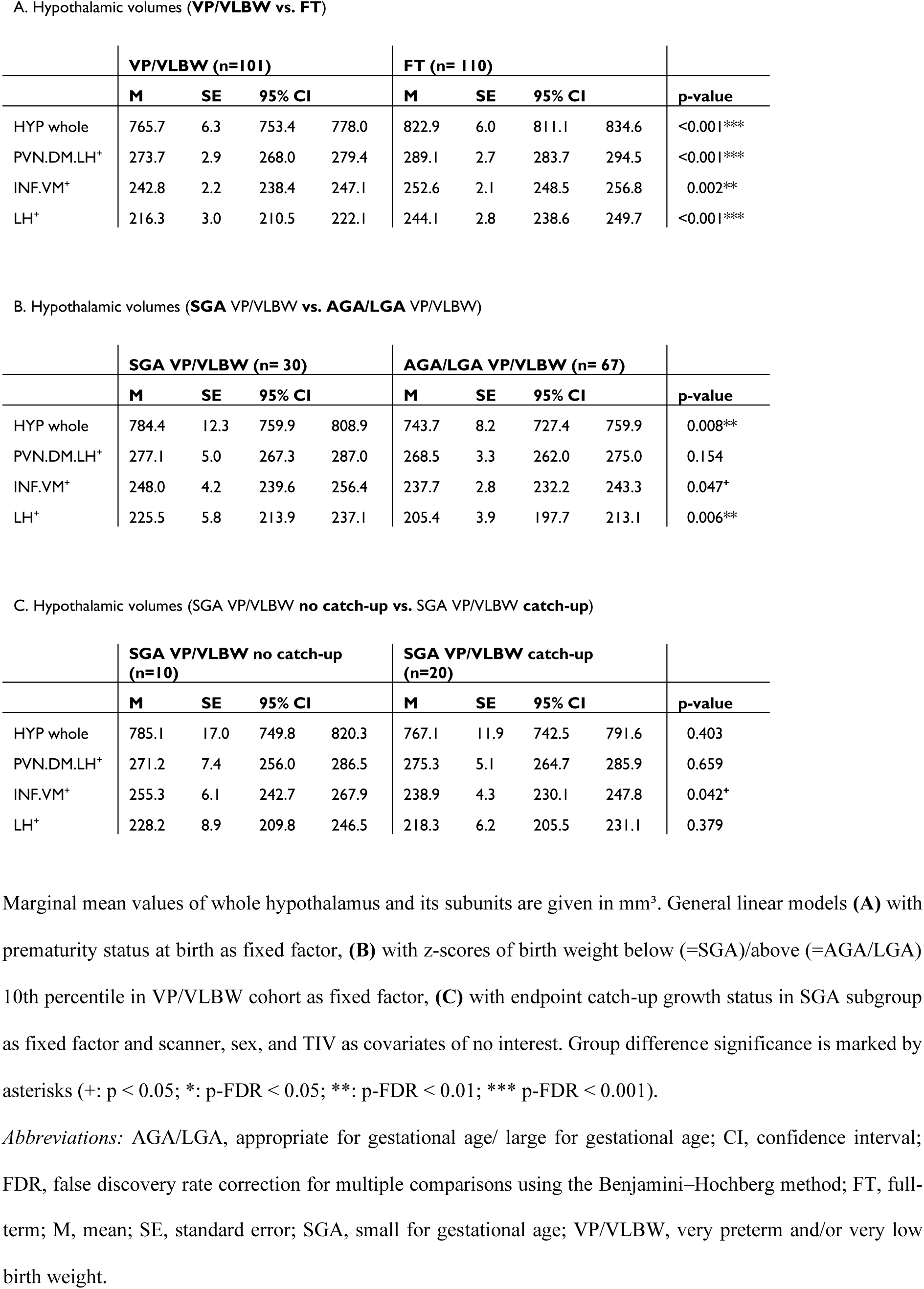
Comparison of hypothalamic volumes for VP/VLBW vs. FT adults, SGA-born vs. AGA/LGA-born VP/VLBW adults and catch-up status yes vs. no in SGA-born VP/VLBW adults.

|  | VP/VLBW (n=101) |  |  |  | FT (n= 110) |  |  |  |  |
| --- | --- | --- | --- | --- | --- | --- | --- | --- | --- |
|  | M | SE | 95% CI |  | M | SE | 95% CI |  | p-value |
| HYP whole | 765.7 | 6.3 | 753.4 | 778.0 | 822.9 | 6.0 | 811.1 | 834.6 | <0.001*** |
| PVN.DM.LH<sup>+</sup> | 273.7 | 2.9 | 268.0 | 279.4 | 289.1 | 2.7 | 283.7 | 294.5 | <0.001*** |
| INF.VM<sup>+</sup> | 242.8 | 2.2 | 238.4 | 247.1 | 252.6 | 2.1 | 248.5 | 256.8 | 0.002** |
| LH<sup>+</sup> | 216.3 | 3.0 | 210.5 | 222.1 | 244.1 | 2.8 | 238.6 | 249.7 | <0.001*** |

B. Hypothalamic volumes (SGA VP/VLBW vs. AGA/LGA VP/VLBW)
|  | SGA VP/VLBW (n= 30) |  |  |  | AGA/LGA VP/VLBW (n= 67) |  |  |  |  |
| --- | --- | --- | --- | --- | --- | --- | --- | --- | --- |
|  | M | SE | 95% CI |  | M | SE | 95% CI |  | p-value |
| HYP whole | 784.4 | 12.3 | 759.9 | 808.9 | 743.7 | 8.2 | 727.4 | 759.9 | 0.008** |
| PVN.DM.LH<sup>+</sup> | 277.1 | 5.0 | 267.3 | 287.0 | 268.5 | 3.3 | 262.0 | 275.0 | 0.154 |
| INF.VM<sup>+</sup> | 248.0 | 4.2 | 239.6 | 256.4 | 237.7 | 2.8 | 232.2 | 243.3 | 0.047<sup>+</sup> |
| LH<sup>+</sup> | 225.5 | 5.8 | 213.9 | 237.1 | 205.4 | 3.9 | 197.7 | 213.1 | 0.006** |

C. Hypothalamic volumes (SGA VP/VLBW no catch-up vs. SGA VP/VLBW catch-up)
|  | SGA VP/VLBW no catch-up<br>(n=10) |  |  |  | SGA VP/VLBW catch-up<br>(n=20) |  |  |  |  |
| --- | --- | --- | --- | --- | --- | --- | --- | --- | --- |
|  | M | SE | 95% CI |  | M | SE | 95% CI |  | p-value |
| HYP whole | 785.1 | 17.0 | 749.8 | 820.3 | 767.1 | 11.9 | 742.5 | 791.6 | 0.403 |
| PVN.DM.LH<sup>+</sup> | 271.2 | 7.4 | 256.0 | 286.5 | 275.3 | 5.1 | 264.7 | 285.9 | 0.659 |
| INF.VM<sup>+</sup> | 255.3 | 6.1 | 242.7 | 267.9 | 238.9 | 4.3 | 230.1 | 247.8 | 0.042<sup>+</sup> |
| LH<sup>+</sup> | 228.2 | 8.9 | 209.8 | 246.5 | 218.3 | 6.2 | 205.5 | 231.1 | 0.379 |

Marginal mean values of whole hypothalamus and its subunits are given in mm<sup>3</sup>. General linear models (A) with prematurity status at birth as fixed factor, (B) with z-scores of birth weight below (=SGA)/above (=AGA/LGA) 10th percentile in VP/VLBW cohort as fixed factor, (C) with endpoint catch-up growth status in SGA subgroup as fixed factor and scanner, sex, and TIV as covariates of no interest. Group difference significance is marked by asterisks (+: p < 0.05; \*: p-FDR < 0.05; \*\*: p-FDR < 0.01; \*\*\* p-FDR < 0.001).
Abbreviations: AGA/LGA, appropriate for gestational age/ large for gestational age; CI, confidence interval; FDR, false discovery rate correction for multiple comparisons using the Benjamini–Hochberg method; FT, full-
term; M, mean; SE, standard error; SGA, small for gestational age; VP/VLBW, very preterm and/or very low birth weight.

To ensure the reliability of the applied hypothalamic segmentation, we compared measured hypothalamic volumes with those of previous hypothalamus parcellation studies (see Supplementary Fig. S2 and Table S2). We found similar hypothalamic volumes for our FT cohort, supporting the reliability of the hypothalamic segmentation.

To test whether hypothalamic volume reductions in VP/VLBW adults are indeed related to preterm birth, we performed partial correlation analyses. We found significant positive associations between GA and volumes of whole hypothalamus (r=0.265, p=0.009), INF.VM^+^ (r=0.247, p=0.016), and LH^+^ (r=0.236, p=0.021) as well as significant negative associations between INTI and volumes of whole hypothalamus (r=-0.296, p=0.004), PVN.DM.LH^+^ (r=- 0.328, p=0.001), and LH^+^ (r=-0.251, p=0.014) (Figure 2C, Supplementary Table S3), suggesting that lower volumes are related with preterm birth.

To test whether hypothalamic volume reductions were related with body weight, we applied both partial correlation and mediation analyses. Concerning correlation analysis, we found at-trend-to-significance correlation (after correction for multiple comparisons) between PVN.DM.LH^+^ volume and adult body weight (r=0.214, p=0.041; Supplementary Table S4). For mediation analysis (Supplementary Fig. S3) the three growth-related hypothalamus subunits, PVN.DM.LH^+^, INF.VM^+^ and LH^+^, were taken as parallel potential mediators of the relation between INTI or GA, respectively, and adult body weight. We found that the PVN.DM.LH^+^ subunit served as a selective mediator of the association between INTI and adult body weight for VP/VLBW adults. These results indicate the relevance of the PVN.DM.LH^+^ subunit for adult body weight after preterm birth.

To test whether VP/VLBW growth trajectories were linked with adult hypothalamic volumes, we performed a trajectory slope analysis in the VP/VLBW group. We identified growth slopes of individuals by adapting a linear regression model to the measured longitudinal body weight z-scores (Figure 2D). Subsequently, change of body weight z-scores from birth to adulthood of regressed body weight trajectories (delta slope) was used for partial correlation analysis with hypothalamic volumes (Figure 2E; Supplementary Table S5). Whole hypothalamus volume (r=0.238, p=0.022), PVN.DM.LH^+^ (r=0.223, p=0.033), and LH^+^ (r=0.216, p=0.039) were at-trend-to-significant positively correlated with growth trajectory slopes, suggesting that growth trajectories link with adult hypothalamic volumes in prematurity.

### Adult hypothalamus, birth weight, and SGA

Next, we studied the association of lower hypothalamic volumes with body weight trajectories in the VP/VLBW group for specific subgroups.

First, we focused on groups of lower birth weight, particularly individuals born SGA. To start with, we tested whether adult hypothalamic volumes were related to birth weight in the VP/VLBW group using a partial correlation approach corrected for sex, scanner, and TIV. We found, unexpectedly, significant *negative* correlations, namely for the whole hypothalamus (r=- 0.262, p=0.012), INF.VM^+^ (r=-0.231, p=0.027), and LH^+^ (r=-0.273; p=0.009) (Supplementary Table S6).

Correspondingly, when comparing VP/VLBW individuals born SGA (n=30) and those born AGA/LGA (n=67), we found larger volumes for the SGA group, significant for whole hypothalamus and LH^+^ and at trend-to-significance for the INF.VM^+^ (Table 2B). To test whether SGA hypothalamic volumes were comparable with those of FT controls, we added FT controls into the general linear model (Figure 3A; Supplementary Table S7) and found indeed that both PVN.DM.LH^+^ and INF.VM^+^ volumes of the SGA group did not differ significantly from those of the FT group.

**Figure 3.**
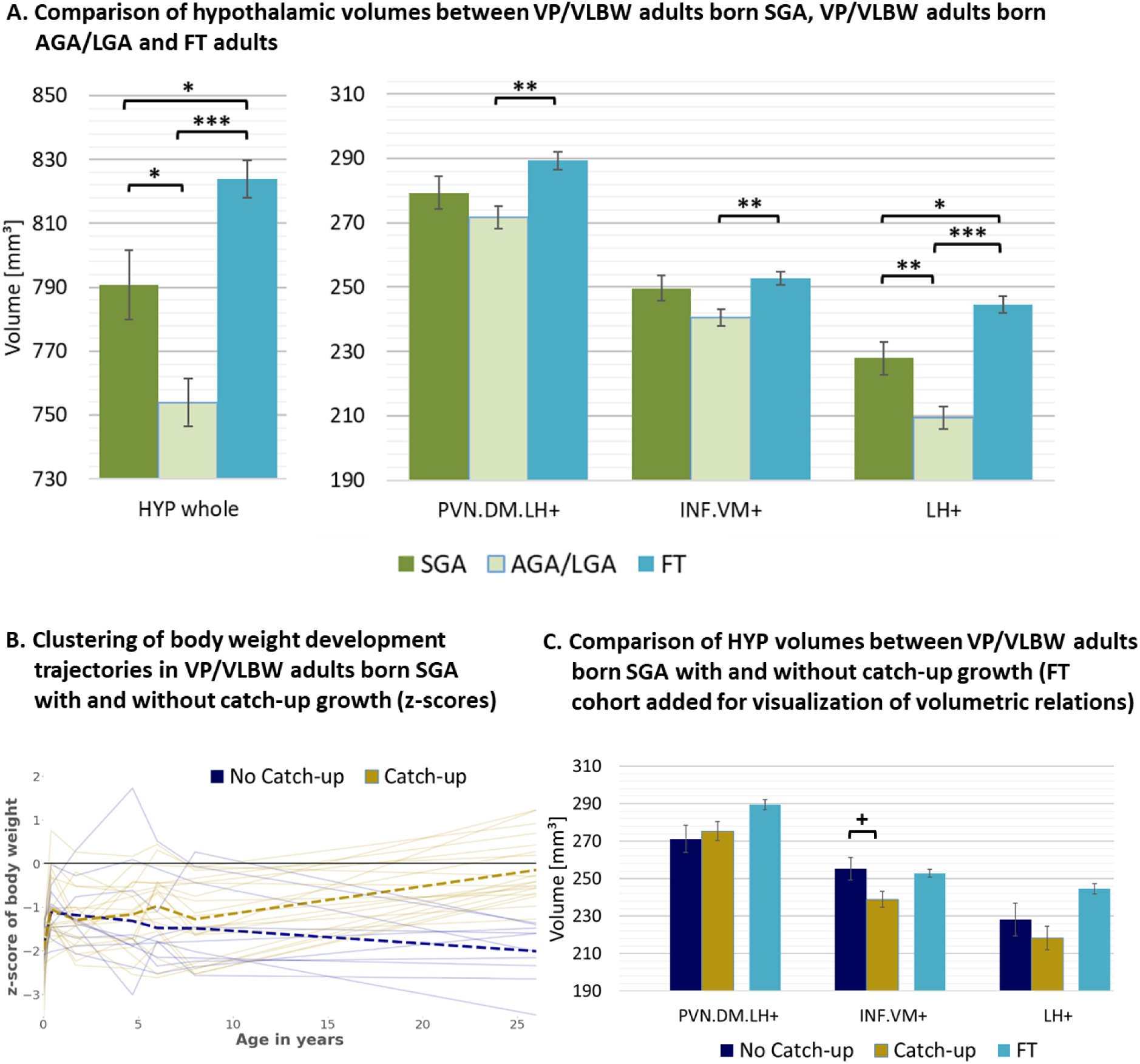
Hypothalamic volumes and birth weight: **(A)** Relatively increased HYP volumes of VP/VLBW adults born SGA compared to those VP/VLBW adults born AGA/LGA. Marginal means of HYP volumes are given in mm³ and are shown as bar plots; error bars indicate SE. Group differences were assessed using a general linear model (fixed factor: SGA, AGA/LGA and FT group differentiation status, covariates of no interest: sex, scanner, TIV). Group difference significance is marked by asterisks (+: p < 0.05; *: p-FDR < 0.05; **: p-FDR < 0.01; *** p-FDR < 0.001). **(B)** Individual body weight development trajectories in the VP/VLBW SGA cohort. Endpoint catch-up growth was assessed successful if SGA individuals exceeded 10th percentile of adult body weight z-score (yellow trajectories) and failed if not (blue trajectories). Dashed bold lines represent mean body weights of both cohorts at each point of time. **(C)** Hypothalamic volumes in catch-up and no catch-up SGA group. Marginal means of HYP subunits are given in mm³ for the SGA cohort comparing catch-up and no catch-up growth and are shown as bar plots; subunit volumes of FT group are added for evaluating volume differences in subgroups relative to FT subjects; error bars indicate SE. Group differences were assessed using a general linear model (fixed factor: endpoint catch-up growth status, covariates of no interest: sex, scanner, TIV). Abbreviations: AGA/LGA, appropriate for gestational age/ large for gestational age; FDR, false discovery rate correction for multiple comparisons using the Benjamini–Hochberg method; FT, full-term; SE, standard error; SGA, small for gestational age; VP/VLBW, very preterm and/or very low birth weight.

Then, we studied body weight development trajectories of VP/VLBW adults born SGA. We asked whether - within this SGA subgroup - hypothalamic volumes differed with respect to individuals’ body weight trajectory after birth, namely for the subgroups of SGA with ‘successful’ (n=20) or ‘failed’ (n=10) endpoint catch-up growth, defined as *adult* body weight z-score being above or below the 10th percentile. Differences between these two groups were not significant for GA (p=0.582), INTI (p= 0.658), and birth weight z-scores (p=0.831), but for adult body weight z-score (p<0.001). Remarkably, the SGA group of failed endpoint catch-up growth had relatively larger volumes than their successful catch-up counterpart for INF.VM^+^ (p<0.05), with volumes in the range of that in the FT group (Figure 3B/C; Table 2C).

### Adult hypothalamus, body weight trajectory into adulthood, and catch-up growth

Second, to study further special subgroups for associations between hypothalamic volumes and body weight trajectories, we focused now on all VP/VLBW individuals who realized long-term catch-up growth (Figure 4). To identify these individuals, we performed a growth trajectory cluster analysis using a k-means algorithm approach. Optimal cluster size (N=4) was determined as a trade-off between maximizing explained variance score via Python (compare Supplementary Fig. S4) and generating functionally explainable body growth trajectory cluster. We found four distinct cluster of body growth development (Figure 4A) which we called “low” (n=32 individuals; 7 were included in the failed endpoint catch-up growth SGA group above), “increasing” (n=18; 13 were included in the successful endpoint catch-up growth SGA group), “decreasing” (n=21) and “average” (n=26) weight trajectory. The “increasing” trajectory group (red color) represented individuals who started with lowest birth weight z-scores (-2.026) in comparison to all other VP/VLBW clustered groups but increased their body weight z-scores during development into adulthood to almost normal; we used this group as a proxy for the long-term catch-up growth group. Using a general linear model with these four clustered groups as fixed factor, we found that the “increasing” group had relatively largest hypothalamic volumes within the VP/VLBW cohort (see Figure 4B), with at-trend-to-significant difference for the LH^+^ subunit (p=0.024), when compared with the “decreasing” group (yellow color). By adding FT cohort as fixed factor into a further general linear model, we found that the “increasing” group’s hypothalamic volumes were not different to those of the FT cohort, particularly not for whole hypothalamus, PVN.DM.LH^+^ and LH^+^ subunits, while for the other groups (i.e., low, decreasing, and average group) whole hypothalamus and LH^+^ volumes were smaller compared to FT (see Supplementary Fig. S5).

**Figure 4.**
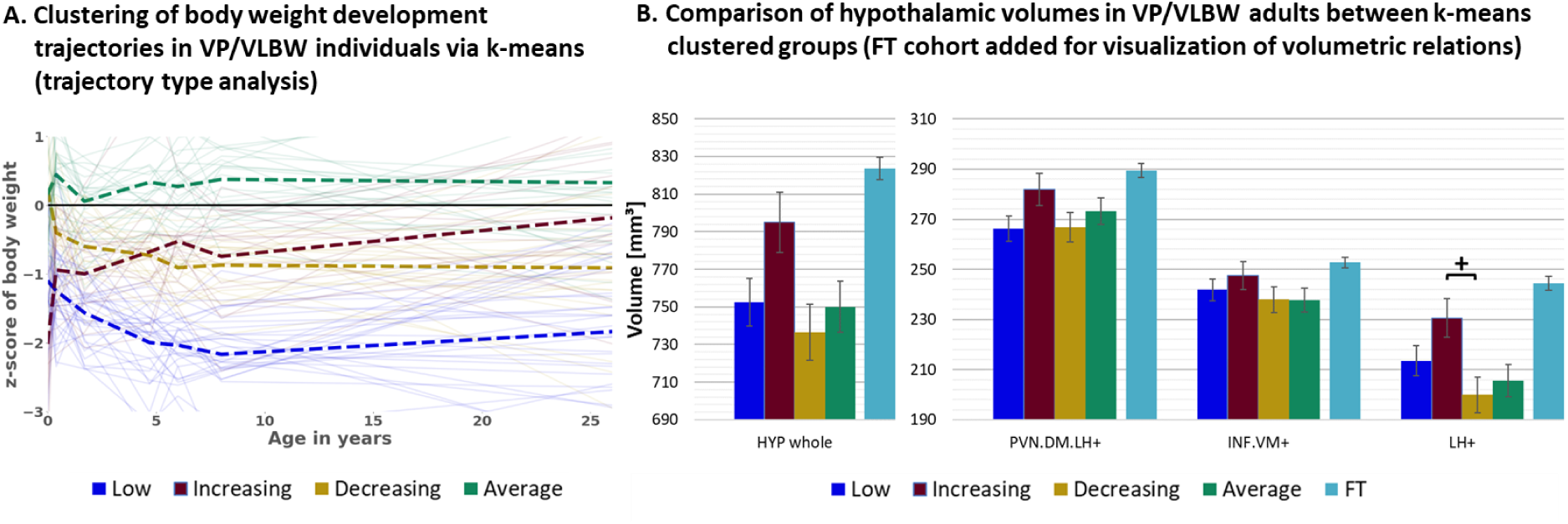
Hypothalamic volumes and body weight trajectories into adulthood: **(A)** Clustering of body weight development trajectories in the VP/VLBW cohort via k-means algorithm (four cluster predefined); k-means grouping: “low” (dark blue), “increasing” (red), “decreasing” (yellow) and “average” (green) (trajectory type analysis). **(B)** Relatively largest HYP volumes of “increasing” group within VP/VLBW cohort. Marginal means of HYP volumes are given in mm³ for k-means clustered groups of VP/VLBW cohort and are shown as bar plots; subunit volumes of FT group (light blue) are added for evaluating volume changes in subgroups relative to FT subjects; error bars indicate SE. Group differences were assessed using a general linear model (fixed factor: four-part variable considering k-means clustered VP/VLBW group differentiation, covariates of no interest: sex, scanner, TIV). Group difference significance is marked by asterisks (+: p < 0.05; *: p-FDR < 0.05; **: p-FDR < 0.01; *** p-FDR < 0.001). Abbreviations: FDR, false discovery rate correction for multiple comparisons using the Benjamini–Hochberg method; FT, full-term; SE, standard error; VP/VLBW, very preterm and/or very low birth weight.

## Discussion

Using T1-weighted MRI, deep-learning-based hypothalamus subunit segmentation, and longitudinal body weight assessment into adulthood, we observed reduced volumes of whole hypothalamus and hypothalamus subunits relevant for growth control in VP/VLBW adults, with volume reductions being distinctively associated with long-term growth trajectories after preterm birth.

### Volumes of growth-related hypothalamus subunits are reduced in VP/VLBW adults and link with growth trajectories into adulthood

Volumes of whole hypothalamus and those of growth-related subunits PVN.DM.LH^+^, INF.VM^+^ and LH^+^ were reduced in VP/VLBW adults compared to FT controls (Fig. 2B). This result was not confounded by sex, scanner, and differences in TIV, as we controlled for these factors. Control of TIV suggests relatively stronger hypothalamic, as compared to general brain volume reduction after preterm birth. Volume reductions of VP/VLBW adults were related to GA and INTI, suggesting that hypothalamic volume reductions were indeed linked with preterm birth (Figure 2C). Hypothalamic volume reductions are in line with long-term volume reductions in other subcortical grey matter structures after preterm birth, such as cholinergic basal forebrain, thalamus or basal ganglia nuclei.^34–37^ Furthermore, hypothalamic volume reductions were associated with both altered adult body weight (Supplementary Fig. S3, Table S4) and growth trajectories in general, namely trajectory slopes (Figure 2E, Supplementary Table S5), indicating that hypothalamic volume reductions are relevant for long-term growth failure of prematurity.

Regarding potential microscopic causes of hypothalamic volume reductions, we speculate that canonical hypoxic-ischemic events-induced pathways of both primary impairment of transient cells, namely pre-oligodendrocytes (Pre-OLs) and subplate neurons (SPNs), and secondary activation of reactive neuroinflammatory cells, such as microglia or astrocytes, are critical.^38–,41^ Pre-OLs are responsible for myelination and axonal maturation of connections, SPNs for the development of the cortical plate including cortical layering and connectivity; these cells are of increased vulnerability to hypoxic-ischemic events, which are typical for preterm birth, their impairment leads amongst others to primary damage of white matter connections, which then results in secondary aberrant grey matter cortical and subcortical development, likely including highly connected hypothalamic nuclei. We also cannot exclude direct hypoxic-ischemic damage on hypothalamic cells. Beyond perinatal brain injuries, it has been further suggested that additional factors, such as malnutrition and maternal, fetal or postnatal stress, do not only alter feeding behavior and growth,^42^ but also hypothalamic development of preterm-born infants.^21, 39, 43–45^

### Distinct long-term growth trajectories and hypothalamus subunits in VP/VLBW adults

#### VP/VLBW adults born SGA have relatively larger hypothalamic volumes that interact with altered growth trajectories into adulthood

We unexpectedly found negative correlations between adult hypothalamic volumes and birth weight in VP/VLBW adults (Supplementary Table S6). Correspondingly, VP/VLBW adults born SGA have relatively larger adult hypothalamic volumes than their AGA/LGA counter-parts (Fig. 3A). Similar results have been reported for pituitary structure, with an increased pituitary size in preterm-born babies being SGA.^46^ Contrary to that, several studies have linked preterm-born infants being SGA or those with intrauterine growth restriction (IUGR) with reduced cortical^47, 48^ and non-hypothalamic subcortical grey matter volumes such as hippocampus, basal ganglia or thalamus nuclei.^49, 50^

To explain relatively increased hypothalamic volumes in VP/VLBW adults born SGA, two non-exclusive scenarios are conceivable: First, hypothalamic volume increases might already be present in infancy or even at birth (as in the case of the pituitary gland) and/or second, they might develop postnatally over time. The first hypothesis is supported by observed hyperactivity of the human hypothalamic-pituitary axis in preterm-born SGA neonates due to hormonal changes (e.g., growth-hormone (GH), IGF-1), suggesting compensatory enlargement of the hypothalamus.^11, 46, 51^ It is also supported by animal studies showing higher cell counts or relative volumes of certain growth-related nuclei in a (non-prematurity) SGA model, potentially due to intrauterine neuroendocrine changes and epigenetic modifications.^52^

The second hypothesis is supported by studies that demonstrated higher vulnerability of grey matter for adverse effects of prematurity on SGA or IUGR infants in comparison to AGA, suggesting a certain degree of perinatal hypothalamic volume reduction.^39, 53^ In this case, the later increased hypothalamic volumes of VP/VLBW born SGA in adulthood could be explained by an altered volumetric hypothalamus development from birth into adulthood.^21^ Notably, long-term hypothalamic development can be affected by prolonged neurogenesis e.g., based on special neural stem cells called tanycytes, which are also supposed to regulate food intake and energy expenditure.^54–56^ These cells are located both along the lateral walls near to growth-related nuclei and at the floor of the third ventricle, namely nearby the INF.VM^+^ sub-unit. Their neurogenesis is controlled e.g., by neurotrophic factors like brain-derived neurotrophic factor (BDNF),^57^ or nutrition and the endocrine system, including metabolic hormones such as leptin, insulin, ghrelin, and IGF-1.^58, 59^ Concentrations of these hormones and molecules are altered in humans after preterm birth,^15, 51, 60^ specifically for SGA-born individuals.^61–63^ Besides prematurity and weight status at birth, growth changes during development are linked with altered ghrelin and leptin levels in humans.^64^ Both hormones affect infundibular and ventromedial nuclei (as parts of INF.VM^+^), main sites of tanycyte neurogenesis.^55^ This might contribute to our findings of VP/VLBW adults born SGA *without* endpoint catch-up growth having relatively larger hypothalamic volumes, particularly for the INF.VM^+^ subunit, than those with catch-up growth (Fig. 3B/C). This result suggests that for the SGA subgroup different hypothalamic nuclei might develop distinctively with respect to body growth development.

While these microscopic mechanisms are speculative with respect to our macroscopic MRI-based findings, they make at least plausible the possibility of distinctively interacting trajectories for body and hypothalamic nuclei growth. These observations demand for further longitudinal investigations of hypothalamic and body weight trajectories. A feasible next step of analysis might be tracking hypothalamic nuclei volumes at birth together with birth weight to decide whether hypothalamic volume reductions in prematurity are already present perinatally or (further) develop postnatally.

#### VP/VLBW adults with body weight catch-up into adulthood show “volumetric catch-up” of hypothalamic volumes to FT adults

By the use of k-means clustering of body weight development trajectories (trajectory type analysis), we identified a special group of VP/VLBW individuals with catch-up growth (Figure 4). Remarkably, hypothalamic nuclei volumes of this VP/VLBW catch-up group were not different to those of the FT group, while the other three VP/VLBW groups had lower hypothalamic volumes for whole hypothalamus and LH^+^ (Supplementary Fig. S5); additionally, LH^+^ volumes of the catch-up group were relatively larger than those of the other VP/VLBW trajectory groups (Fig. 4B). Therefore, relatively larger hypothalamic volumes, particularly for subunits including the lateral hypothalamus, seem to be associated with more favorable long-term growth trajectories in VP/VLBW adults. This observation is in line with findings that volumetric increases of hypothalamic nuclei are linked with altered hypothalamic regulation of body weight.^65^ Furthermore, it matches results in humans that revealed a positive association of general brain growth with optimized early nutrition^66^ and body growth.^67^ Given the link between favorable long-term growth trajectory and growth-related hypothalamic nuclei volumes, we suggest that hypothalamus mapping after birth might have some potential to predict risks for long-term growth failure. Future studies of the infant hypothalamus are necessary to test this suggestion, with implications for both optimized nutrition^68^ and neuroprotective/neurorestorative interventions^39^ to prevent potential aberrant hypothalamic development.

### Strengths and limitations

Strengths of our study are the large sample size, enhancing power and generalizability of our findings. Homogenous mean across VP/VLBW and FT groups excludes confounding age effects. Additionally, hypothalamus segmentation quality was optimized by using high-resolution T1-weighted images.

Nevertheless, our results should be viewed as conservative estimates of the true group differences including hypothalamic volumes, as our sample is biased towards VP/VLBW adults with less severe neonatal complications and thus adult impairments. VP/VLBW adults with more neonatal complications or functional impairments had an increased probability not to participate in the study due to MRI exclusion criteria or to reject MRI screening. Nevertheless, the sample was still representative of the full Bavarian Longitudinal Study cohort in terms of GA and BW.^30^

One of the major limitations of our study is the subunit-focused parcellation methodology of the hypothalamus segmentation algorithm by Billot et al..^28^ As their subunit definition is bound to anatomical landmarks, therefore aggregating several hypothalamic nuclei per subunit, it is challenging to allocate differences in subunit volumes to distinct growth-related hypothalamic nucleus volumes and hence their functional implications. Additionally, MRI brain scans were only available at age 26, whereas a longitudinal measurement of hypothalamic volumes during development would be required to address several relevant issues, such as separating cause from consequence regarding altered hypothalamic structure or revealing hypothalamic structure interdependency with body growth.

In conclusion, our results demonstrate an association of preterm birth with growth control-related hypothalamus subunits and long-term growth failure after preterm birth.

## Data Availability

The data that support the findings of this study are available from the corresponding author upon reasonable request.

## Acknowledgements

We thank all current and former members of the Bavarian Longitudinal Study Group who contributed to general study organization, recruitment, data collection and management as well as subsequent analyses, including (in alphabetical order): Barbara Busch, Stephan Czeschka, Claudia Grünzinger, Christian Koch, Diana Kurze, Sonja Perk, Andrea Schreier, Antje Strasser, Julia Trummer, and Eva van Rossum. We are grateful to the staff of the Department of Neuroradiology in Munich and the Department of Radiology in Bonn for their help in data collection. Most importantly, we thank all our study participants and their families for their efforts to take part in this study.

## Funding

This study was supported by the Deutsche Forschungsgemeinschaft (BA 6370/2-1 to C.S.), the German Federal Ministry of Education and Science (BMBF 01ER0801 to P.B. and D.W., BMBF 01ER0803 to C.S.) and the Kommission für Klinische Forschung, Technische Universität München (KKF 8700000474 to D.M.H.).

## Competing interests

The authors report no competing interests.

## Abbreviations

AGA/LGA: appropriate/large for gestational age
BW: birth weight
FT: full-term
GA: gestational age
HYP: hypothalamus
INTI: intensity of neonatal treatment index
SGA: small for gestational age
TIV: total intracranial volume
VP/VLBW: very preterm/very low birth weight.

## Supplementary Materials and methods

### Participants

This study assessed a geographically defined whole population sample of neonatal at-risk very preterm and/or very low birth weight (<32 weeks of gestation and/or <1500g) individuals (VP/VLBW) and healthy full-term controls (FT). Sample data were collected as part of the Bavarian Longitudinal Study, of which a more detailed description can be found elsewhere.^1, 2^ Briefly, all individuals were born between Jan 1^st^, 1985, and March 31^st^, 1986, in a defined region of Southern Bavaria and were followed from birth into adulthood. The VP/VLBW cohort initially consisted of 682 individuals of whom 260 participated in the 26-year follow-up assessment, including measurement of body growth parameters. Of the initial 916 FT controls from the same obstetric hospitals alive at 6 years, 350 were randomly selected within the stratification variables of sex and family socioeconomic status as being comparable to the VP/VLBW cohort. 229 of them participated in the 26-year follow-up assessment. For subsequent brain imaging at age 26, all individuals were screened for MR-related exclusion criteria including (self-reported) claustrophobia, inability to lie still for > 30min, unstable medical conditions (e.g., severe asthma), epilepsy, tinnitus, pregnancy, non-removable MRI-incompatible metal implants, and a history of severe central nervous system (CNS) trauma or disease that would impair further analysis of the data. The most frequent reason not to perform the MRI exam, however, was lack of motivation. The remaining 101 VP/VLBW individuals and 111 FT controls underwent MRI scan. The MRI examinations took place at two sites: The Department of Neuroradiology, Klinikum rechts der Isar, Technical University of Munich (n=146), and the Department of Radiology, University Hospital of Bonn (n=66). For a detailed flowchart of participants through the study see Supplement Fig. S1 and^3^.

The study was carried out in accordance with the Declaration of Helsinki and was approved by the local ethics committee of the Klinikum rechts der Isar and the University Hospital Bonn. All study participants gave written informed consent. They received travel expenses and payment for participation.

### Gestation and medical treatment

Gestational age (GA) was estimated from maternal reports on the first day of the last menstrual period and from serial ultrasounds during pregnancy. In cases, in which the two measures differed by more than 2 weeks, clinical assessment at birth with the Dubowitz method was applied.^4^ To estimate medical impairments at birth, Intensity of Neonatal Treatment Index (INTI) was calculated via daily assessments of care level, respiratory support, feeding dependency, and neurological status (mobility, muscle tone, and neurological excitability). Each of these six variables was scored on a 4-point rating scale (0–3) using the method of Casaer and Eggermont^5^ (see Supplement Table S1 for a description of the variables). INTI was computed as the mean score of daily ratings during the first 10 days of life or until a stable clinical state was reached (total daily scores <3 for 3 consecutive days), depending on which occurred first, ranging from 0 (best state) to 18 (worst state).

### Body growth

Body weight measurements were undertaken at birth and during follow-up visits at five and 20 months corrected for prematurity, and at 56 months, 6, 8 and 26 years of chronological age by specially trained research nurses. They used predefined protocols with weighing on standard scales in underwear only.^6^ For growth analysis, we transformed body weight measurements from [g]/[kg] into z-scores relative to the exact age of each participant, typically used for the description of longitudinal changes of growth status.^7, 8^ In detail, a neonatal reference was applied for calculation of “birth weight for gestational age z-scores” (referred to as “birth weight z-scores” in our study). We used Voigt’s neonatal population database comprising 2.3 million live and still singleton births in Germany from 1995 to 2000 with a GA from 20 to 43 weeks,^9, 10^ allowing for sex-specific weight percentiles to be calculated. For calculation of body weight z-scores at all other ages a reference was applied comprising percentiles of body weight in children and adolescents including 17.147 males and 17.275 females, aged 0-18 years, evaluated from different regional German studies.^11^ For ages greater 18 years, the authors additionally integrated percentile data from the German “Mikrozensus”^12^ into their reference.

For all analyses focusing on variables regarding body growth restricted to the VP/VLBW cohort, four individuals were excluded as adult body weight at age 26 was missing (remaining number of individuals of VP/VLBW cohort for body growth analyses: N=97).

#### SGA status and catch-up growth

“Small for gestational age” (SGA) versus “appropriate/large for gestational age” (AGA/LGA) was determined depending on birth weight z-scores being below (equivalent to z-scores < -1.282) versus above the 10th percentile, respectively.^9, 13^ Within the SGA subcohort, we defined successful endpoint catch-up growth as an adult body weight z-score at age 26 above the 10th percentile. In doing so, we concentrated on long-term body growth development as opposed to short-term evaluations of catch-up growth within the first two years of life as mostly performed in the literature.^14–17^ This allowed us to better account for the additional role of long-term growth in metabolic and cardiovascular outcome after preterm birth.^18, 19^

#### Body growth trajectory analysis

To analyze the relationship between hypothalamic volumes and body growth trajectories, we performed both trajectory slope and trajectory type analysis using Python version 3.7.10 and especially the “scikit-learn” package, a machine learning focused Python library.

Firstly, for trajectory slope analysis we assessed body growth development restricted to the VP/VLBW group. Linear regression of body weight z-scores (non-interpolated) at birth, ages 5, 20, 56 months and 6, 8 and 26 years was performed to receive regressed body weight trajectories. Partial correlations were used to investigate the associations between change of body weight z-score from birth until adulthood of regressed body weight trajectories (delta slope) and hypothalamic volumes corrected for sex, scanner, and total intracranial volume (TIV; sum of segmented grey and white matter brain volumes and cerebrospinal fluid partitions).

Secondly, for trajectory type analysis, we revealed growth trajectory types in the VP/VLBW group by a clustering approach. In particular, we clustered body growth development trajectories via k-means algorithm in Python (sklearn.cluster.KMeans) after approximating missing data points via linear interpolation (number of missing data points can be deduced from Table 1 with regards to missing samples from maximum N=97 VP/VLBW, N=110 FT at every stage of longitudinal weight analysis). The k-means algorithm is based on minimizing within-cluster sum of squared distances between each data point and the centroids.^20–22^ The number of times the k-means algorithm was run with different centroid seeds as default initialization was N=100. After trajectory clustering, hypothalamic volumes were compared across cluster using general linear models as described above.

### MRI data acquisition

MRI data acquisition (see^23^) was performed at Klinikum rechts der Isar, Technical University of Munich, and Bonn University Hospital on Philips Achieva 3 T systems or Philips Ingenia 3T systems using an 8-channel SENSE head coil. Subject distribution among scanners was: Bonn Achieva 3T: 5 VP/VLBW, 11 FT; Bonn Ingenia 3T: 33 VP/VLBW, 17 FT; Munich Achieva 3T: 60 VP/VLBW, 65 FT; Munich Ingenia 3T: 3 VP/VLBW, 17 FT. Across all scanners sequence parameters were kept identical. Scanners were checked regularly to provide optimal scanning conditions and MRI physicists at the University Hospital Bonn and Klinikum rechts der Isar regularly scanned imaging phantoms to ensure within-scanner signal stability over time. Signal-to-noise ratio was not significantly different between scanners (one-way analysis of variance with factor “scanner- ID” [Bonn 1, Bonn 2, Munich 1, Munich 2]; F(3,182)=1.84, p=0.11). A high-resolution T1-weighted 3D magnetization prepared rapid acquisition gradient echo (MPRAGE) sequence (TI=1,300ms, TR=7.7ms, TE=3.9ms, flip angle=15°; 180 sagittal slices, FOV=256×256×180mm, reconstruction matrix=256×256; reconstructed isotropic voxel size=1mm³) was acquired. All images were visually inspected for artefacts. In our study, to account for possible confounds by scanner differences, MRI data analyses included scanner dummy variables as covariates of no interest.

### Hypothalamus segmentation

T1-weighted MRI scans in Nifti-format were processed by using the freely available FreeSurfer image analysis suite (http://surfer.nmr.mgh.harvard.edu/). Particularly, the version of Free-Surfer 7.1.1 includes a deep convolutional neural network tool of Billot et al.^24^ that enables for automated segmentation of the hypothalamus, including subsegment parcellation. Billot and colleagues trained this neural network on thirty-seven T1-weighted, manually labelled, and augmented MRI scans. A state-of-the-art 3D U-net model^25^ served as a basis for the architecture of the network. As a result, this hypothalamus delineation approach exceeded former approaches based on multi-atlas segmentation or deep learning-based algorithms,^26^ and it was comparable to expert intra-rater precision.^24^ We applied the algorithm to our dataset of T1- weighted MRI scans.

To focus on hypothalamic nuclei involved in growth control, we mapped subsegments from the segmentation algorithm that contained nuclei relevant for growth control onto *three so-called subunits of growth control, namely PVN.DM.LH^+^, INF.VM^+^ and LH^+^*. The PVN.DM.LH^+^ subunit is the aggregate of two subsegments of the segmentation of Billot et al.,^24^ namely the anterior-superior and superior tubular subsegment (see Fig. 1), and these two subsegments cover the preoptic area, the paraventricular nucleus (PVN), the dorsomedial nucleus (DM), and parts of the lateral hypothalamus (LH), with the latter three being involved in growth control and therefore defining the name of the subunit. INF.VM^+^ is identical to the inferior tubular subsegment, which comprises - amongst other nuclei - the growth control-related infundibular (INF) and ventromedial (VM) nuclei. LH^+^ matches the posterior subsegment in Billot et al.,^24^ including the mammillary bodies, parts of the tuberomammillary nucleus and, critically, of the growth control-related lateral hypothalamus (LH). The final anterior-inferior subsegment (suprachiasmatic nucleus and parts of the supraoptic nucleus) was excluded from further analysis because it does not contain any growth-related nuclei.

All subunits stated in the analysis already consider volumes of bilateral hypothalamus.

### Statistical analysis

Statistical analyses were performed using SPSS version 27 (IBM SPSS Statistics).

#### Analyses of group differences and correlation analyses

Regarding demographical characteristics, group differences between VP/VLBW and FT cohorts were assessed using chi-square tests (sex) and two-sample t-tests (age, GA). To test whether both hypothalamic volumes or body weights are altered in prematurity, general linear models were used (dependent variable: hypothalamic volumes and body weight, respectively; fixed factor: status of prematurity at birth; covariates: sex and additionally only for hypothalamic volume changes: scanner, TIV). Age was not included as a covariate in our analyses, as VP/VLBW subjects and FT controls were not significantly different in mean age at scanning of 26 years. Partial correlation analysis, restricted to the VP/VLBW group and corrected for sex, scanner and TIV, was used to investigate the associations between hypothalamic volumes and variables of preterm birth i.e., GA and INTI, respectively. Regarding variables related to body growth (birth weight z-scores and adult weight) similar correlation analyses were applied.

#### Mediation analysis

To assess potential mediation effects of hypothalamic volumes regarding the association of variables of prematurity (i.e., INTI and GA, respectively) with adult body weight, a mediation analysis restricted to the VP/VLBW cohort was performed using the PROCESS toolbox (version 3.5) of SPSS.^27^ In the mediation model, INTI and GA were entered as causal variables, respectively, adult body weight as the outcome variable, and volumes of PVN.DM.LH^+^, INF.VM^+^ and LH^+^ subunits were introduced simultaneously as potential parallel mediator variables (covariates of no interest: sex, scanner, and TIV). Path coefficients for total effect, direct effect and indirect effect were estimated using (unstandardized) regression coefficients from multiple regression analyses, and statistical significance of the indirect effect was tested using a nonparametric bootstrap approach (with 5000 repetitions) to obtain 95% confidence intervals. We calculated p-values for indirect effects based on 95% confidence intervals, SE and estimated effect as described by Altman and Bland.^28^

#### Statistical thresholds

Statistical significance was set at p <0.05; all tests were two-sided. Tests were corrected for multiple comparisons for false discovery rate (FDR) according to the Benjamini-Hochberg procedure.^29^

## Supplementary Figures

**Figure S1:**
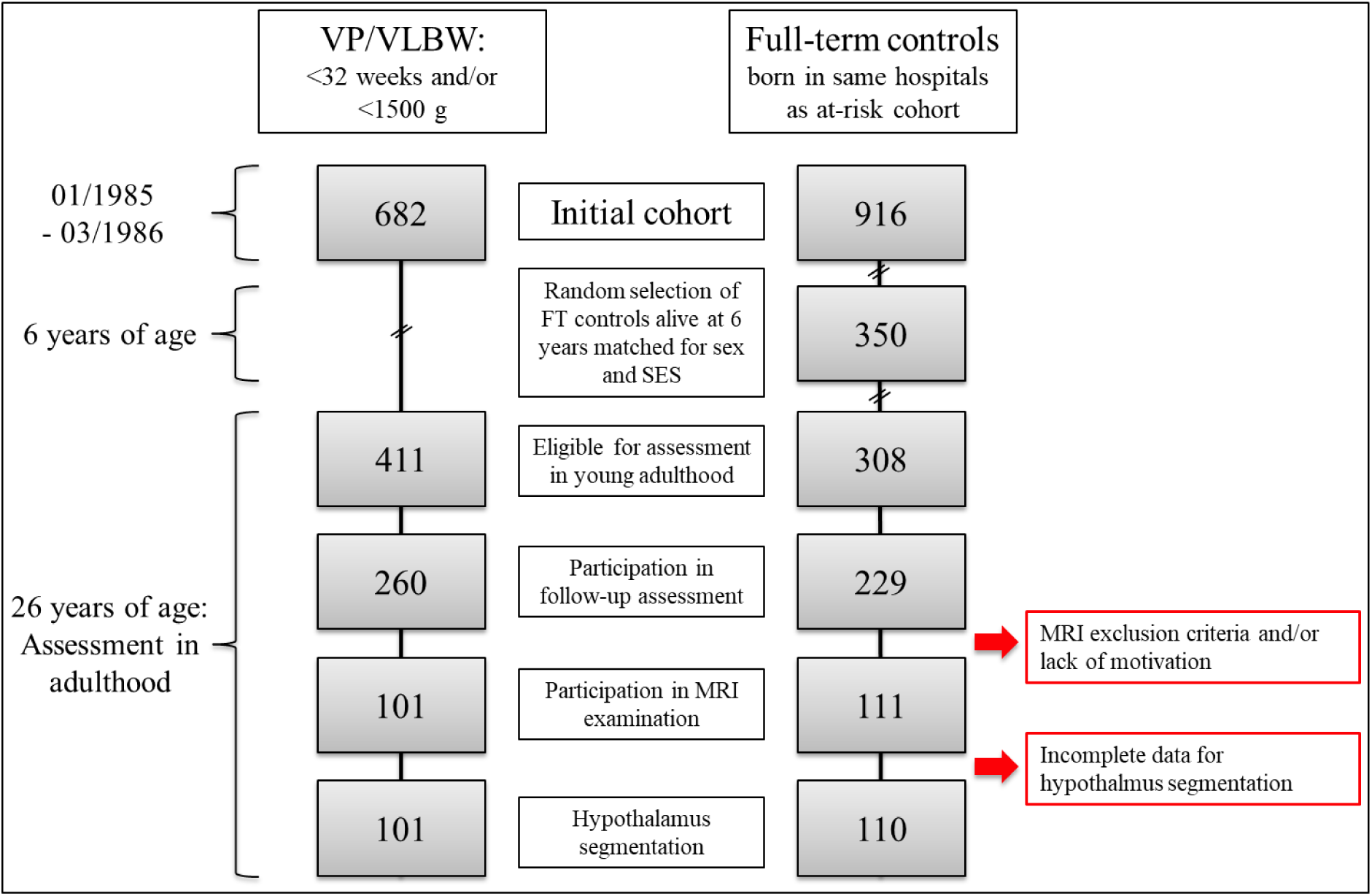
Participants of the Bavarian Longitudinal Study. Flowchart of the participants of the Bavarian Longitudinal Study. Abbreviations: MRI, magnetic resonance imaging; SES, socioeconomic status; VP/VLBW, very preterm and/or very low birth weight.

**Figure S2:**
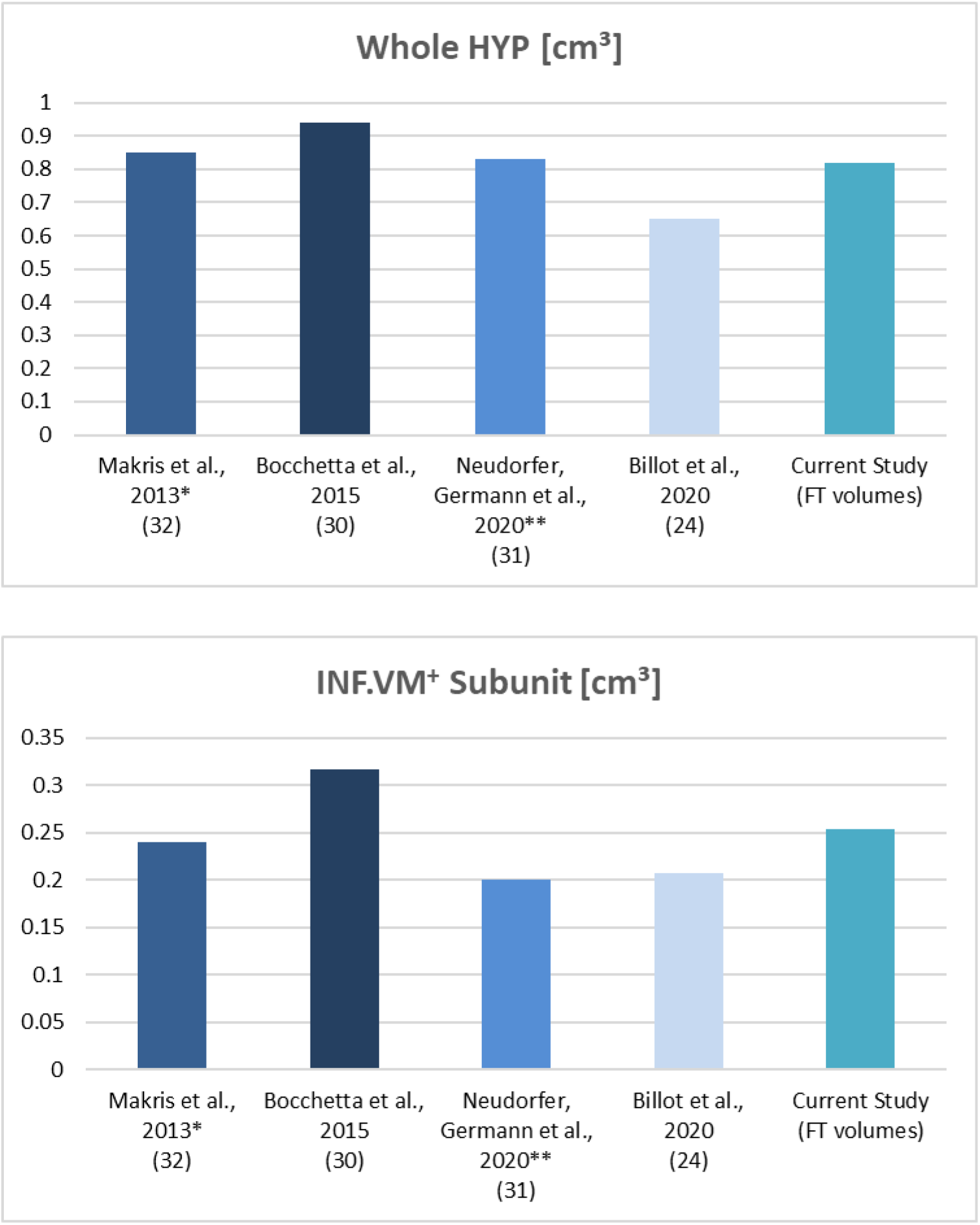
Reliability control analysis of segmented whole HYP and representative functional subunit (INF.VM^+^) volumes via comparison to HYP studies in literature. Comparison of manually or (semi-)automated segmented hypothalamic volumes (in cm³) in research literature for whole hypothalamus (upper line) and hypothalamus subunits with representative visualization of INV.VM^+^ subunit (lower line). With regards to subunit comparisons, studies were selected which provided similar hypothalamus subunit parcellation methodology. If available, segmentation results of healthy controls were taken for comparison (also compare Supplement Table S2). Additional remarks: The INF.VM^+^ (inferior tubular) subunit comprises, according to the Billot et al.^24^ segmentation algorithm, the infundibular nucleus, the ventromedial nucleus, the lateral tubular nucleus, and parts of the tuberomammillary and the supraoptic nuclei; the lateral tubular nucleus was not assigned to the INF.VM^+^ subunit in Bocchetta et al.^30^ and Neudorfer, Germann et al.^31^ and the tuberomammillary nucleus not assigned to the INF.VM^+^ subunit in Makris et al.^32^ and Bocchetta et al..^30^ *HYP volumes = (male + female volumes)/2 (mean of provided values). **Summation of single nuclear volumes (as parcellation on nuclear level is available in Neudorfer, Germann et al.).^31^ Abbreviations: FT, full-term; HYP, hypothalamus.

**Figure S3:**
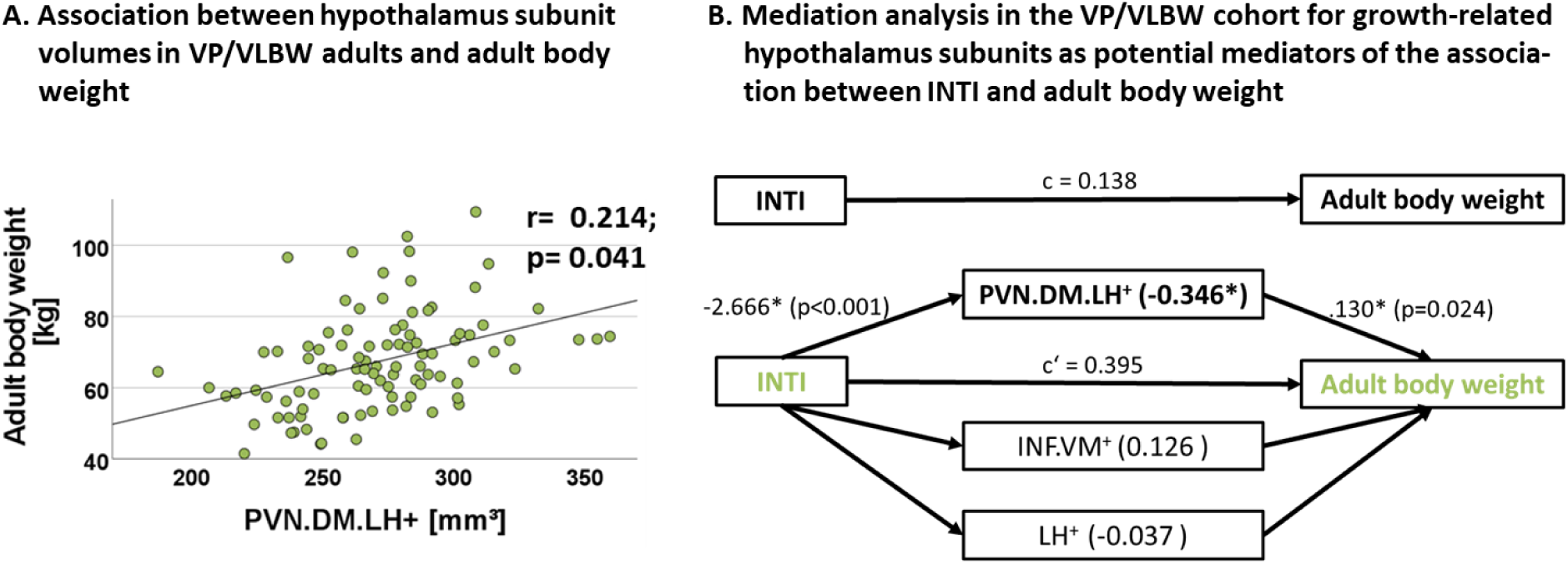
VP/VLBW adults: adult body weight and PVN.DM.LH^+^. **(A)** Association between hypothalamic volumes and adult body weight in the VP/VLBW cohort (compare also Supplement Table S4). Scatterplot shows relationship between adult body weight and PVN.DM.LH^+^ subunit volumes. Linear regression line and regression coefficient of partial regression analysis are added. **(B)** The PVN.DM.LH^+^ subunit volume mediates the association between medical complications at birth (represented by INTI as causal variable) and adult body weight (in kg). A path diagram is shown in order to illustrate the results of the mediation analyses restricted to the VP/VLBW cohort. The regression model was corrected for sex, scanner and TIV. Body weight related hypothalamus subunits were introduced as potential mediators and the PVN.DM.LH^+^ subunit volume yielded a significant indirect effect (ab= -0.346 ± 0.174; bootstrapped 95% CI: -0.738 to -0.075; p= 0.040).^27^ All other subunits did not show significant mediation effects. The figure includes the following standardized regression coefficients: c, the total effect of INTI on adult body weight; c’, the direct effect of INTI on adult body weight when adjusting for the potential mediating variables. Significant regression coefficients (p < 0.05) are marked with an asterisk. For GA as causal variable, we did not find a similar mediation effect. Abbreviations: INTI, intensity of neonatal treatment; VP/VLBW, very preterm and/or very low birth weight.

**Figure S4:**
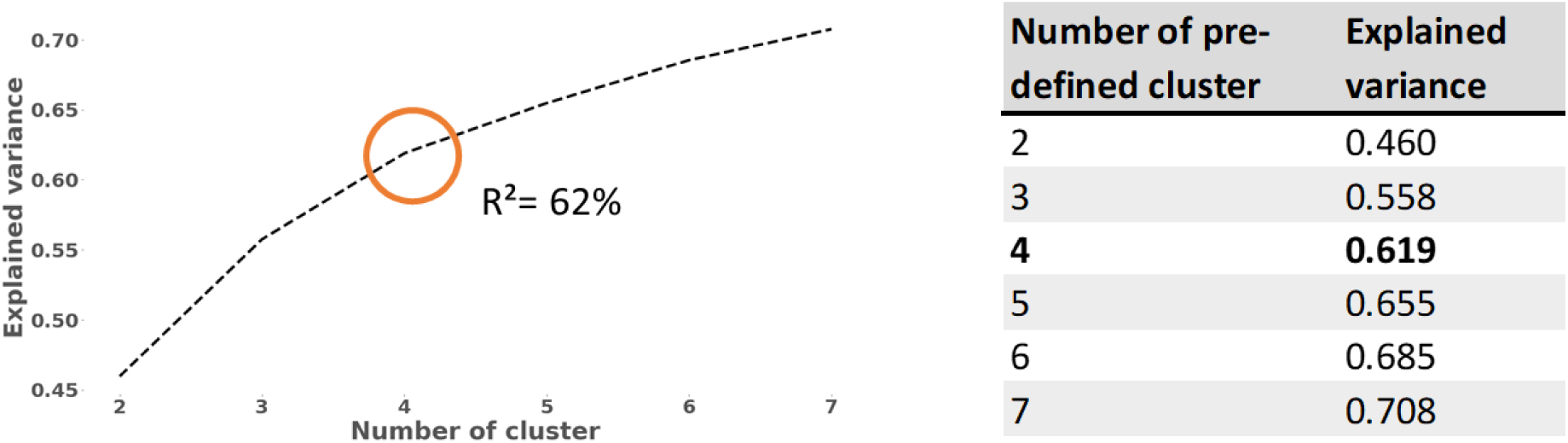
K-means cluster analysis of body weight trajectories: differences in explained variance regression score with varying number of predefined cluster. ***Left.*** Graph of explained variance regression score with varying number of predefined cluster for clustering of body weight development trajectories in the VP/VLBW cohort. The explained variance regression score (sklearn.metrics.explained_variance_score) is calculated as *explained_variance(y, y*)= 1- Var(y - y*) / Var(y)*, where *y – y\** describes the pointwise calculation of the gap between the individual trajectory under observation (y) and the correspondent clustered trajectory (y*). Var as variance represents the square of the standard deviation. The explained variance regression score is ≤ 1, with 1 being the optimum. ***Right.*** Absolute values of explained variance regression scores depending on number of predefined cluster. Abbreviations: VP/VLBW, very preterm and/or very low birth weight.

**Figure S5:**
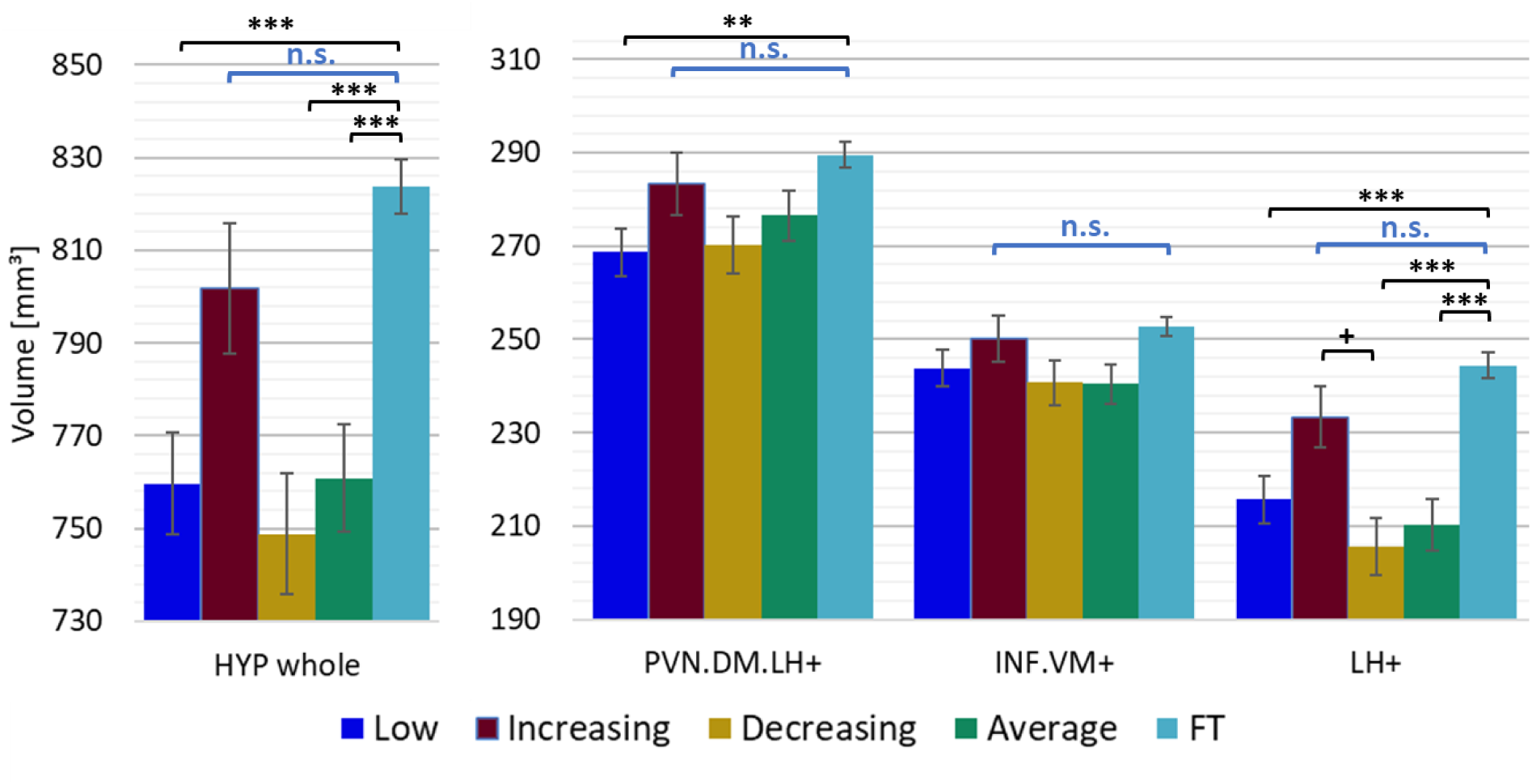
Comparison of hypothalamic volumes between k-means clustered groups of VP/VLBW adults and adult FT cohort. Marginal means of hypothalamic volumes are given in mm³ for FT cohort and k-means clustered groups of VP/VLBW cohort and are shown as bar plots; error bars indicate SE. Group differences were assessed using a general linear model (fixed factor: five-part variable considering k-means clustered VP/VLBW and FT group differentiation; covariates of no interest: sex, scanners, TIV). Group difference significance is marked by asterisks (+: p < 0.05; *: p-FDR < 0.05; **: p-FDR < 0.01; *** p-FDR < 0.001). Abbreviations: FDR, false discovery rate correction for multiple comparisons using the Benjamini–Hochberg method; FT, full-term; n.s., not significant; SE, standard error; VP/VLBW, very preterm and/or very low birth weight.

## Supplementary Tables

**Table S1.**
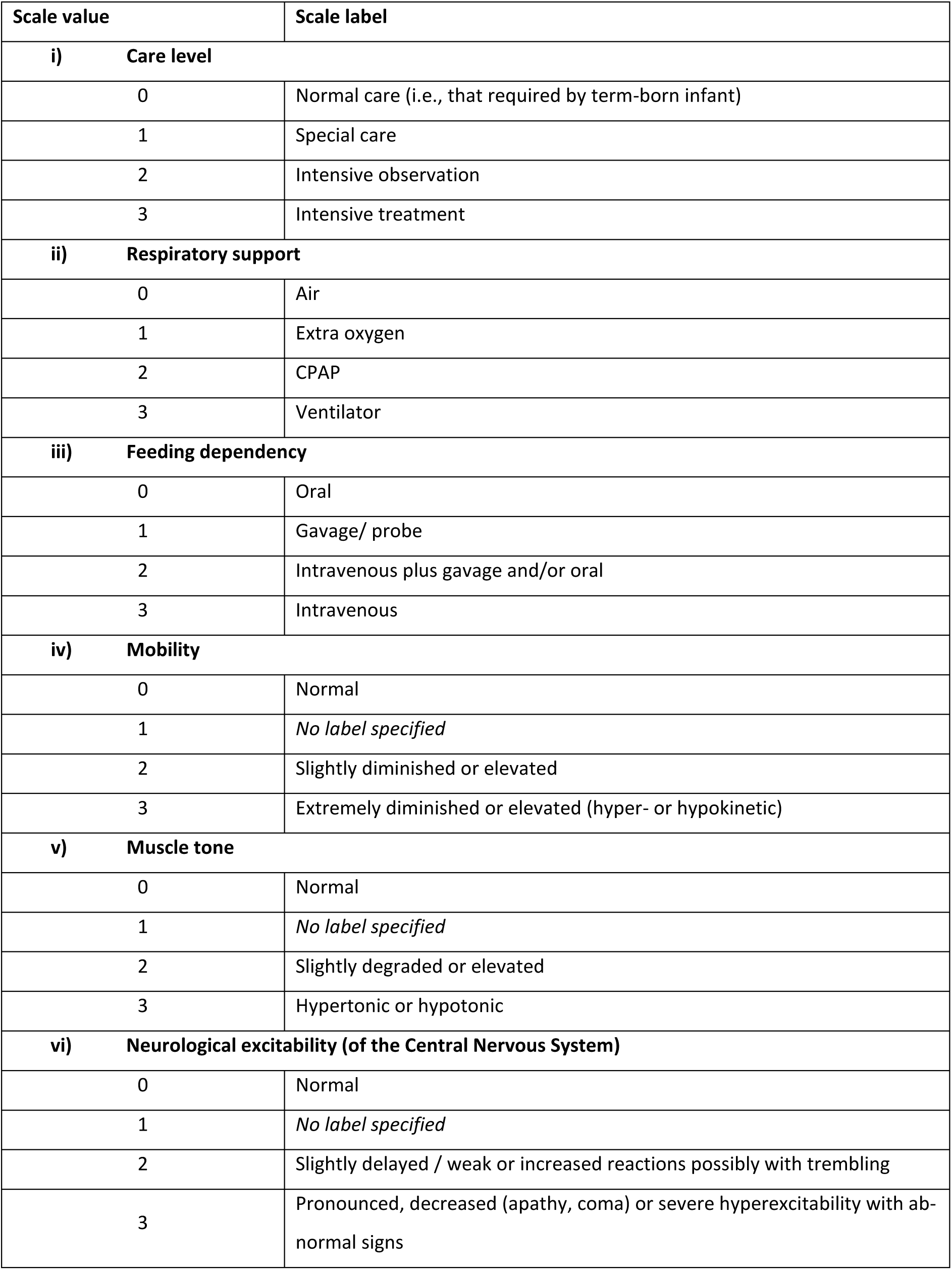
Description of the six variables that comprise the INTI. Abbreviations: CPAP, continuous positive airway pressure; INTI, intensity of neonatal treatment index.

**Table S2:**
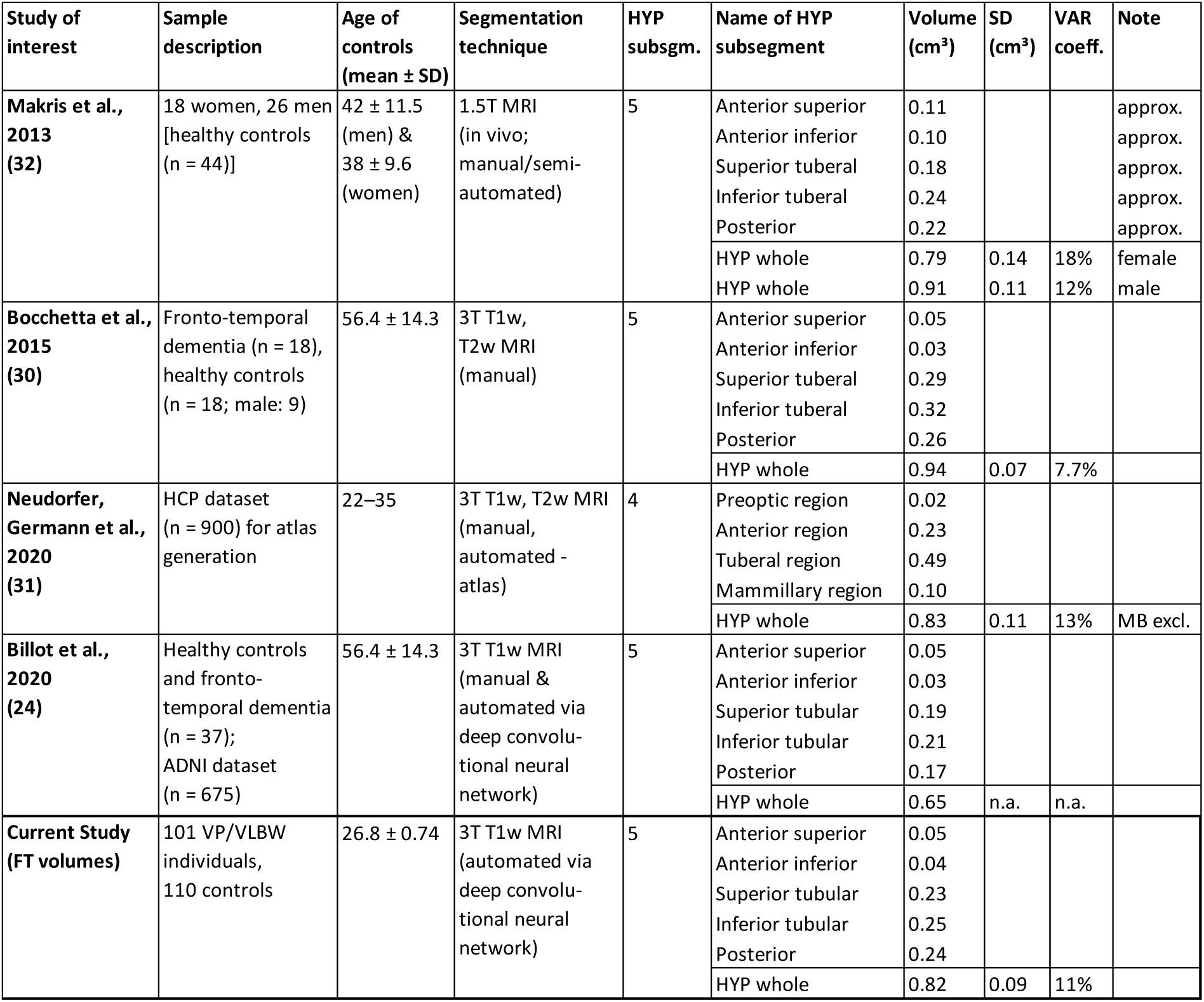
Hypothalamus parcellation methodologies and segmentation results in literature. Comparison of manually or (semi-)automated segmented hypothalamic volumes (in cm³) in research literature, their parcellation methodology and segmentation approach for whole hypothalamus and hypothalamus subunits. For visualization of hypothalamic volume comparisons see Supplement Fig. S2. More examples of whole hypothalamus segmentation, parcellation methodologies and volumetric data in literature can be found in^33–35^, from where some of the data in the table is also extracted from. Abbreviations: approx., approximated; ADNI, Alzheimer’s Disease Neuroimaging Initiative; HCP, Human Connectome Project; MB excl., mammillary bodies excluded; SD, standard deviation; subsgm., subsegment; VAR coeff., variation coefficient; VP/VLBW, very preterm and/or very low birth weight.

**Table S3:**
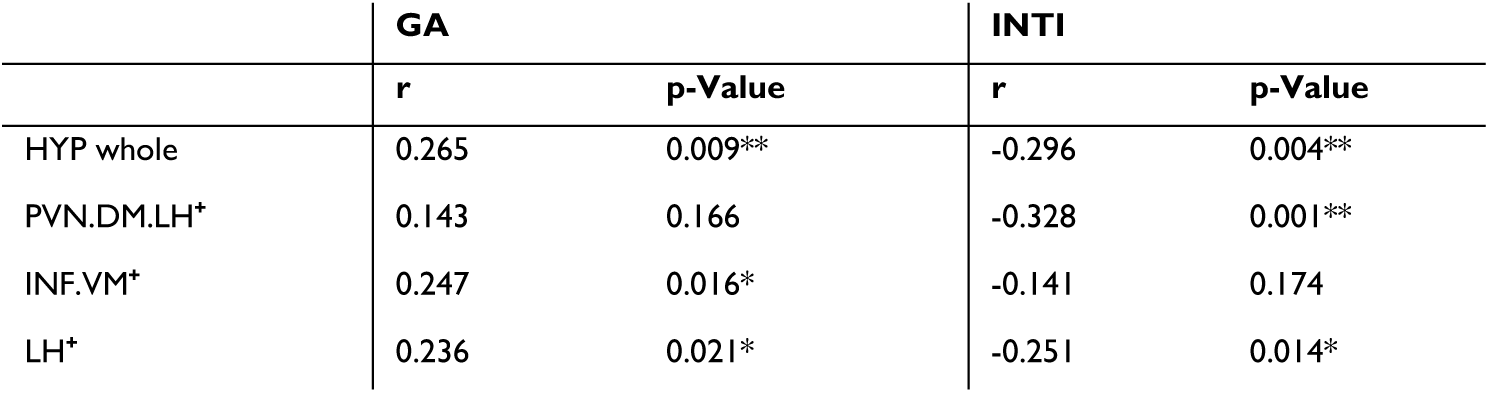
Relationship between hypothalamic volumes in VP/VLBW adults and variables of preterm birth. Correlation coefficients from partial correlation analyses in the VP/VLBW sample are given. TIV, scanner, and sex served as covariates. Group difference significance is marked by asterisks (+: p < 0.05; *: p-FDR < 0.05; **: p-FDR < 0.01; *** p-FDR < 0.001). Abbreviations: FDR, false discovery rate correction for multiple comparisons using the Benjamini–Hochberg method; GA, gestational age; INTI, intensity of neonatal treatment; VP/VLBW, very preterm and/or very low birth weight.

**Table S4:** Relationship between hypothalamic volumes in VP/VLBW adults and adult body weight. Correlation coefficients from partial correlation analyses in the VP/VLBW sample are given. TIV, scanner, and sex served as covariates. Group difference significance is marked by asterisks (+: p < 0.05; *: p-FDR < 0.05; **: p-FDR < 0.01; *** p-FDR < 0.001). Abbreviations: FDR, false discovery rate correction for multiple comparisons using the Benjamini–Hochberg method; VP/VLBW, very preterm and/or very low birth weight.

|  | Adult body weight |  |
| --- | --- | --- |
|  | r | p-Value |
| HYP whole | 0.164 | 0.118 |
| PVN.DM.LH<sup>+</sup> | 0.214 | 0.041<sup>+</sup> |
| INF.VM<sup>+</sup> | -0.035 | 0.740 |
| LH<sup>+</sup> | 0.098 | 0.352 |

**Table S5:**
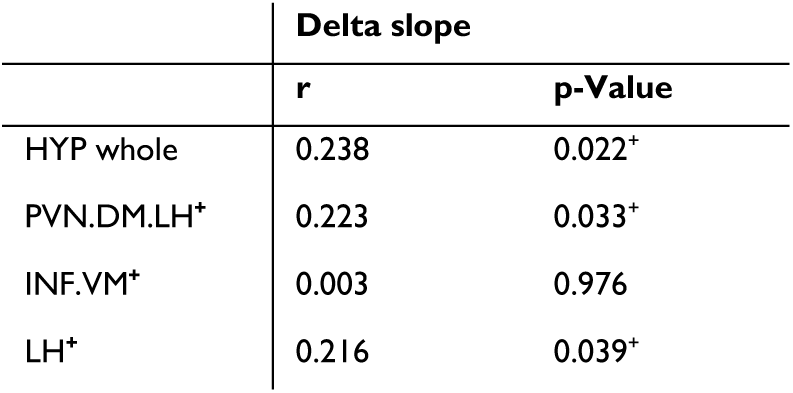
Relationship between hypothalamic volumes in VP/VLBW adults and delta slope of body weight development trajectories. Correlation coefficients from partial correlation analyses in the VP/VLBW sample are given. Delta slope of individual body weight development trajectories is calculated via change of body weight z-score from birth until adulthood of regressed body weight trajectories. TIV, scanner, and sex served as covariates. Group difference significance is marked by asterisks (+: p < 0.05; *: p-FDR < 0.05; **: p-FDR < 0.01; *** p-FDR < 0.001). Abbreviations: FDR, false discovery rate correction for multiple comparisons using the Benjamini–Hochberg method; VP/VLBW, very preterm and/or very low birth weight.

|  | Delta slope |  |
| --- | --- | --- |
|  | r | p-Value |
| HYP whole | 0.238 | 0.022<sup>+</sup> |
| PVN.DM.LH<sup>+</sup> | 0.223 | 0.033<sup>+</sup> |
| INF.VM<sup>+</sup> | 0.003 | 0.976 |
| LH<sup>+</sup> | 0.216 | 0.039<sup>+</sup> |

**Table S6:** Relationship between hypothalamic volumes in VP/VLBW adults and birth weight z-scores. Correlation coefficients from partial correlation analyses in the VP/VLBW sample are given. TIV, scanner, and sex served as covariates. Group difference significance is marked by asterisks (+: p < 0.05; *: p-FDR < 0.05; **: p-FDR < 0.01; *** p-FDR < 0.001). Abbreviations: FDR, false discovery rate correction for multiple comparisons using the Benjamini–Hochberg method; VP/VLBW, very preterm and/or very low birth weight.

|  | Birth weight z-score |  |
| --- | --- | --- |
|  | r | p-Value |
| HYP whole | -0.262 | 0.012* |
| PVN.DM.LH<sup>+</sup> | -0.082 | 0.438 |
| INF.VM<sup>+</sup> | -0.231 | 0.027* |
| LH<sup>+</sup> | -0.273 | 0.009** |

**Table S7:** Group comparison of hypothalamic volumes with regards to birth weight and status of preterm birth (VP/VLBW adults born SGA vs. VP/VLBW adults born AGA/LGA vs. adult FT) Marginal mean values of whole hypothalamus and its subunits are given in mm³. General linear model with SGA, AGA/LGA and FT group differentiation status at birth as fixed factor. Scanner, sex, and TIV served as covariates of no interest. Group difference significance is marked by asterisks (+: p < 0.05; *: p-FDR < 0.05; **: p-FDR < 0.01; *** p-FDR < 0.001). Abbreviations: AGA/LGA, appropriate for gestational age/ large for gestational age; CI, confidence interval; FDR, false discovery rate correction for multiple comparisons using the Benjamini–Hochberg method; FT, full-term; M, mean; SE, standard error; SGA, small for gestational age; VP/VLBW, very preterm and/or very low birth weight.

|  | VP/VLBW SGA (n=30) |  |  |  | VP/VLBW AGA/LGA (n= 67) |  |  |  |  |
| --- | --- | --- | --- | --- | --- | --- | --- | --- | --- |
|  | M | SE | 95% CI |  | M | SE | 95% CI |  | p-value |
| HYP whole (mm³) | 790.8 | 10.9 | 769.3 | 812.2 | 753.9 | 7.5 | 739.1 | 768.8 | 0.016* |
| PVN.DM.LH+ (mm³) | 279.3 | 5.1 | 269.3 | 289.3 | 271.6 | 3.5 | 264.7 | 278.6 | 0.627 |
| INF.VM+ (mm³) | 249.6 | 3.9 | 242.0 | 257.3 | 240.5 | 2.7 | 235.2 | 245.8 | 0.150 |
| LH+ (mm³) | 228.0 | 5.1 | 218.0 | 238.0 | 209.4 | 3.5 | 202.4 | 216.3 | 0.008** |

|  | VP/VLBW SGA (n=30) |  |  |  | FT (n=110) |  |  |  |  |
| --- | --- | --- | --- | --- | --- | --- | --- | --- | --- |
|  | M | SE | 95% CI |  | M | SE | 95% CI |  | p-value |
| HYP whole (mm³) | 790.8 | 10.9 | 769.3 | 812.2 | 823.8 | 5.9 | 812.3 | 835.4 | 0.027* |
| PVN.DM.LH+ (mm³) | 279.3 | 5.1 | 269.3 | 289.3 | 289.4 | 2.7 | 284.0 | 294.8 | 0.262 |
| INF.VM+ (mm³) | 249.6 | 3.9 | 242.0 | 257.3 | 252.8 | 2.1 | 248.7 | 257.0 | 1.000 |
| LH+ (mm³) | 228.0 | 5.1 | 218.0 | 238.0 | 244.6 | 2.7 | 239.2 | 250.0 | 0.015* |

|  | VP/VLBW AGA/LGA (n= 67) |  |  |  | FT (n=110) |  |  |  |  |
| --- | --- | --- | --- | --- | --- | --- | --- | --- | --- |
|  | M | SE | 95% CI |  | M | SE | 95% CI |  | p-value |
| HYP whole (mm³) | 753.9 | 7.5 | 739.1 | 768.8 | 823.8 | 5.9 | 812.3 | 835.4 | <0.001*** |
| PVN.DM.LH+ (mm³) | 271.6 | 3.5 | 264.7 | 278.6 | 289.4 | 2.7 | 284.0 | 294.8 | 0.001** |
| INF.VM+ (mm³) | 240.5 | 2.7 | 235.2 | 245.8 | 252.8 | 2.1 | 248.7 | 257.0 | 0.002** |
| LH+ (mm³) | 209.4 | 3.5 | 202.4 | 216.3 | 244.6 | 2.7 | 239.2 | 250.0 | <0.001*** |

## Notes

### Competing Interest Statement

The authors have declared no competing interest.

### Author Declarations

The study was carried out in accordance with the Declaration of Helsinki and was approved by the local ethics committee of the Klinikum rechts der Isar and the University Hospital Bonn.

## References

1. Chawanpaiboon S, Vogel JP, Moller A-B, et al. Global, regional, and national estimates of levels of preterm birth in 2014: a systematic review and modelling analysis. Lancet Glob Health. 2019;7(1):e37–e46. doi:10.1016/S2214-109X(18)30451-0

2. Markopoulou P, Papanikolaou E, Analytis A, Zoumakis E, Siahanidou T. Preterm Birth as a Risk Factor for Metabolic Syndrome and Cardiovascular Disease in Adult Life: A Systematic Review and Meta-Analysis. J Pediatr. 2019;210:69–80.e5. doi:10.1016/j.jpeds.2019.02.041

3. Abitbol CL, Rodriguez MM. The long-term renal and cardiovascular consequences of prematurity. Nat Rev Nephrol. 2012;8(5):265–274. doi:10.1038/nrneph.2012.38

4. Wolke D, Johnson S, Mendonça M. The Life Course Consequences of Very Preterm Birth. Annu Rev Dev Psychol. 2019;1(1):69–92. doi:10.1146/annurev-devpsych-121318-084804

5. Eves R, Mendonça M, Baumann N, et al. Association of Very Preterm Birth or Very Low Birth Weight With Intelligence in Adulthood: An Individual Participant Data Meta-analysis. JAMA Pediatr. 2021. doi:10.1001/jamapediatrics.2021.1058

6. Euser AM, Wit CC de, Finken MJJ, Rijken M, Wit JM. Growth of preterm born children. Horm Res. 2008;70(6):319–328. doi:10.1159/000161862

7. Ford GW, Doyle LW, Davis NM, Callanan C. Very low birth weight and growth into adolescence. Arch Pediatr Adolesc Med. 2000;154(8):778–784. doi:10.1001/archpedi.154.8.778

8. Lee SM, Kim N, Namgung R, Park M, Park K, Jeon J. Prediction of Postnatal Growth Failure among Very Low Birth Weight Infants. Sci Rep. 2018;8(1):3729. doi:10.1038/s41598-018-21647-9

9. Geisler I, Rausch TK, Göpel W, Spiegler J. Extremely and very preterm-born children <1500 g show different weight development in childhood compared to their peers. Acta Paediatr. 2021. doi:10.1111/apa.15785

10. Boguszewski MCS, Cardoso-Demartini AdA. Management of endocrine disease: Growth and growth hormone therapy in short children born preterm. Eur J Endocrinol. 2017;176(3):R111–R122. doi:10.1530/EJE-16-0482

11. Saenger P, Czernichow P, Hughes I, Reiter EO. Small for gestational age: short stature and beyond. Endocr Rev. 2007;28(2):219–251. doi:10.1210/er.2006-0039

12. Lapillonne A, Griffin IJ. Feeding preterm infants today for later metabolic and cardiovascular outcomes. J Pediatr. 2013;162(3 Suppl):S7–16. doi:10.1016/j.jpeds.2012.11.048

13. Ordóñez-Díaz MD, Pérez-Navero JL, Flores-Rojas K, et al. Prematurity With Extrauterine Growth Restriction Increases the Risk of Higher Levels of Glucose, Low-Grade of Inflammation and Hypertension in Prepubertal Children. Front Pediatr. 2020;8:180. doi:10.3389/fped.2020.00180

14. Ruys CA, Hollanders JJ, Bröring T, et al. Early-life growth of preterm infants and its impact on neurodevelopment. Pediatr Res. 2019;85(3):283–292. doi:10.1038/s41390-018-0139-0

15. Möllers LS, Yousuf EI, Hamatschek C, et al. Metabolic-endocrine disruption due to preterm birth impacts growth, body composition, and neonatal outcome. Pediatr Res. 2021. doi:10.1038/s41390-021-01566-8

16. Yeo GSH, Heisler LK. Unraveling the brain regulation of appetite: lessons from genetics. Nat Neurosci. 2012;15(10):1343–1349. doi:10.1038/nn.3211

17. Morton GJ, Meek TH, Schwartz MW. Neurobiology of food intake in health and disease. Nat Rev Neurosci. 2014;15(6):367–378. doi:10.1038/nrn3745

18. Swaab DF. The Human Hypothalamus : Basic and Clinical Aspects, Part I: Basic and Clinical Aspects - Part I: Nuclei of the Human Hypothalamus. Elsevier; 2003-2004.

19. Makris N, Swaab DF, van der Kouwe A, et al. Volumetric parcellation methodology of the human hypothalamus in neuroimaging: normative data and sex differences. Neuroimage. 2013;69:1–10. doi:10.1016/j.neuroimage.2012.12.008

20. Koutcherov Y, Mai JK, Ashwell KWS, Paxinos G. Organization of human hypothalamus in fetal development. The Journal of Comparative Neurology. 2002;446(4):301–324. doi:10.1002/cne.10175

21. Sominsky L, Jasoni CL, Twigg HR, Spencer SJ. Hormonal and nutritional regulation of postnatal hypothalamic development. J Endocrinol. 2018;237(2):R47–R64. doi:10.1530/JOE-17-0722

22. Andermann ML, Lowell BB. Toward a Wiring Diagram Understanding of Appetite Control. Neuron. 2017;95(4):757–778. doi:10.1016/j.neuron.2017.06.014

23. Thomas K, Beyer F, Lewe G, et al. Higher body mass index is linked to altered hypothalamic microstructure. Sci Rep. 2019;9(1):17373. doi:10.1038/s41598-019-53578-4

24. Florent V, Baroncini M, Jissendi-Tchofo P, et al. Hypothalamic Structural and Functional Imbalances in Anorexia Nervosa. Neuroendocrinology. 2020;110(6):552–562. doi:10.1159/000503147

25. Saper CB. 15 - Hypothalamus. In: Paxinos G, ed. The Human Nervous System. Academic Press; 1990:389–413.

26. Neudorfer C, Germann J, Elias GJB, Gramer R, Boutet A, Lozano AM. A high-resolution in vivo magnetic resonance imaging atlas of the human hypothalamic region. Sci Data. 2020;7(1):305. doi:10.1038/s41597-020-00644-6

27. Spindler M, Özyurt J, Thiel CM. Automated diffusion-based parcellation of the hypothalamus reveals subunit-specific associations with obesity. Sci Rep. 2020;10(1):22238. doi:10.1038/s41598-020-79289-9

28. Billot B, Bocchetta M, Todd E, Dalca AV, Rohrer JD, Iglesias JE. Automated segmentation of the hypothalamus and associated subunits in brain MRI. Neuroimage. 2020;223:117287. doi:10.1016/j.neuroimage.2020.117287

29. Wolke D, Meyer R. Cognitive status, language attainment, and prereading skills of 6-year-old very preterm children and their peers: the Bavarian Longitudinal Study. Dev Med Child Neurol. 1999;41(2):94–109. doi:10.1017/s0012162299000201

30. Schmitz-Koep B, Zimmermann J, Menegaux A, et al. Decreased amygdala volume in adults after premature birth. Sci Rep. 2021;11(1):5403. doi:10.1038/s41598-021-84906-2

31. Gutbrod T, Wolke D, Soehne B, Ohrt B, Riegel K. Effects of gestation and birth weight on the growth and development of very low birthweight small for gestational age infants: a matched group comparison. Arch Dis Child Fetal Neonatal Ed. 2000;82(3):F208–14. doi:10.1136/fn.82.3.f208

32. Onis M de, Habicht JP. Anthropometric reference data for international use: recommendations from a World Health Organization Expert Committee. Am J Clin Nutr. 1996;64(4):650–658. doi:10.1093/ajcn/64.4.650

33. Benjamini Y, Hochberg Y. Controlling the False Discovery Rate: A Practical and Powerful Approach to Multiple Testing. Journal of the Royal Statistical Society. Series B (Methodological). 1995;57(1):289–300. http://www.jstor.org/stable/2346101

34. Ball G, Boardman JP, Rueckert D, et al. The effect of preterm birth on thalamic and cortical development. Cereb Cortex. 2012;22(5):1016–1024. doi:10.1093/cercor/bhr176

35. Nosarti C, Nam KW, Walshe M, et al. Preterm birth and structural brain alterations in early adulthood. Neuroimage Clin. 2014;6:180–191. doi:10.1016/j.nicl.2014.08.005

36. Loh WY, Anderson PJ, Cheong JLY, et al. Longitudinal growth of the basal ganglia and thalamus in very preterm children. Brain Imaging Behav. 2020;14(4):998–1011. doi:10.1007/s11682-019-00057-z

37. Grothe MJ, Scheef L, Bäuml J, et al. Reduced Cholinergic Basal Forebrain Integrity Links Neonatal Complications and Adult Cognitive Deficits After Premature Birth. Biol Psychiatry. 2017;82(2):119–126. doi:10.1016/j.biopsych.2016.12.008

38. Salmaso N, Jablonska B, Scafidi J, Vaccarino FM, Gallo V. Neurobiology of premature brain injury. Nat Neurosci. 2014;17(3):341–346. doi:10.1038/nn.3604

39. Volpe JJ. Dysmaturation of Premature Brain: Importance, Cellular Mechanisms, and Potential Interventions. Pediatr Neurol. 2019;95:42–66. doi:10.1016/j.pediatrneurol.2019.02.016

40. Reemst K, Noctor SC, Lucassen PJ, Hol EM. The Indispensable Roles of Microglia and Astrocytes during Brain Development. Front Hum Neurosci. 2016;10:566. doi:10.3389/fnhum.2016.00566

41. Hoerder-Suabedissen A, Molnár Z. Development, evolution and pathology of neocortical subplate neurons. Nat Rev Neurosci. 2015;16(3):133–146. doi:10.1038/nrn3915

42. Maniam J, Morris MJ. The link between stress and feeding behaviour. Neuropharmacology. 2012;63(1):97–110. doi:10.1016/j.neuropharm.2012.04.017

43. Lammertink F, Vinkers CH, Tataranno ML, Benders, Manon J N L. Premature Birth and Developmental Programming: Mechanisms of Resilience and Vulnerability. Front Psychiatry. 2020;11:531571. doi:10.3389/fpsyt.2020.531571

44. Howland MA, Sandman CA, Glynn LM. Developmental origins of the human hypothalamic-pituitary-adrenal axis. Expert Rev Endocrinol Metab. 2017;12(5):321–339. doi:10.1080/17446651.2017.1356222

45. Finken MJJ, van der Voorn B, Hollanders JJ, et al. Programming of the Hypothalamus-Pituitary-Adrenal Axis by Very Preterm Birth. Ann Nutr Metab. 2017;70(3):170–174. doi:10.1159/000456040

46. Kiortsis D, Xydis V, Drougia AG, et al. The height of the pituitary in preterm infants during the first 2 years of life: an MRI study. Neuroradiology. 2004;46(3):224–226. doi:10.1007/s00234-003-1126-6

47. Tolsa CB, Zimine S, Warfield SK, et al. Early alteration of structural and functional brain development in premature infants born with intrauterine growth restriction. Pediatr Res. 2004;56(1):132–138. doi:10.1203/01.PDR.0000128983.54614.7E

48. Miller SL, Huppi PS, Mallard C. The consequences of fetal growth restriction on brain structure and neurodevelopmental outcome. J Physiol. 2016;594(4):807–823. doi:10.1113/JP271402

49. Lodygensky GA, Seghier ML, Warfield SK, et al. Intrauterine growth restriction affects the preterm infant’s hippocampus. Pediatr Res. 2008;63(4):438–443. doi:10.1203/PDR.0b013e318165c005

50. Padilla N, Junqué C, Figueras F, et al. Differential vulnerability of gray matter and white matter to intrauterine growth restriction in preterm infants at 12 months corrected age. Brain Res. 2014;1545:1–11. doi:10.1016/j.brainres.2013.12.007

51. Kajantie E. Insulin-like growth factor (IGF)-I, IGF binding protein (IGFBP)-3, phosphoisoforms of IGFBP-1 and postnatal growth in very-low-birth-weight infants. Horm Res. 2003;60 Suppl 3:124–130. doi:10.1159/000074513

52. Plagemann A, Harder T, Rake A, Melchior K, Rohde W, Dörner G. Hypothalamic nuclei are malformed in weanling offspring of low protein malnourished rat dams. J Nutr. 2000;130(10):2582–2589. doi:10.1093/jn/130.10.2582

53. Vinall J, Grunau RE, Brant R, et al. Slower postnatal growth is associated with delayed cerebral cortical maturation in preterm newborns. Sci Transl Med. 2013;5(168):168ra8. doi:10.1126/sci-translmed.3004666

54. Pellegrino G, Trubert C, Terrien J, et al. A comparative study of the neural stem cell niche in the adult hypothalamus of human, mouse, rat and gray mouse lemur (Microcebus murinus). The Journal of Comparative Neurology. 2018;526(9):1419–1443. doi:10.1002/cne.24376

55. Bolborea M, Dale N. Hypothalamic tanycytes: potential roles in the control of feeding and energy balance. Trends Neurosci. 2013;36(2):91–100. doi:10.1016/j.tins.2012.12.008

56. Bolborea M, Langlet F. What is the physiological role of hypothalamic tanycytes in metabolism? Am J Physiol Regul Integr Comp Physiol. 2021;320(6):R994–R1003. doi:10.1152/aj-pregu.00296.2020

57. Pencea V, Bingaman KD, Wiegand SJ, Luskin MB. Infusion of brain-derived neurotrophic factor into the lateral ventricle of the adult rat leads to new neurons in the parenchyma of the striatum, septum, thalamus, and hypothalamus. J Neurosci. 2001;21(17):6706–6717. doi:10.1523/JNEURO-SCI.21-17-06706.2001

58. Chaker Z, George C, Petrovska M, et al. Hypothalamic neurogenesis persists in the aging brain and is controlled by energy-sensing IGF-I pathway. Neurobiol Aging. 2016;41:64–72. doi:10.1016/j.neurobiolaging.2016.02.008

59. Desai M, Li T, Ross MG. Fetal hypothalamic neuroprogenitor cell culture: preferential differentiation paths induced by leptin and insulin. Endocrinology. 2011;152(8):3192–3201. doi:10.1210/en.2010-1217

60. Krey FC, Stocchero BA, Creutzberg KC, et al. Neurotrophic Factor Levels in Preterm Infants: A Systematic Review and Meta-Analysis. Front Neurol. 2021;12:643576. doi:10.3389/fneur.2021.643576

61. Martos-Moreno GA, Barrios V, Sáenz de Pipaón M, et al. Influence of prematurity and growth restriction on the adipokine profile, IGF1, and ghrelin levels in cord blood: relationship with glucose metabolism. Eur J Endocrinol. 2009;161(3):381–389. doi:10.1530/EJE-09-0193

62. Jaquet D, Leger J, Levy-Marchal C, Oury JF, Czernichow P. Ontogeny of leptin in human fetuses and newborns: effect of intrauterine growth retardation on serum leptin concentrations. J Clin Endocrinol Metab. 1998;83(4):1243–1246. doi:10.1210/jcem.83.4.4731

63. Finken MJJ, van der Steen M, Smeets CCJ, et al. Children Born Small for Gestational Age: Differential Diagnosis, Molecular Genetic Evaluation, and Implications. Endocr Rev. 2018;39(6):851–894. doi:10.1210/er.2018-00083

64. Monti V, Carlson JJ, Hunt SC, Adams TD. Relationship of ghrelin and leptin hormones with body mass index and waist circumference in a random sample of adults. J Am Diet Assoc. 2006;106(6):822–8; quiz 829-30. doi:10.1016/j.jada.2006.03.015

65. Le TM, Liao D-L, Ide J, et al. The interrelationship of body mass index with gray matter volume and resting-state functional connectivity of the hypothalamus. Int J Obes (Lond). 2020;44(5):1097–1107. doi:10.1038/s41366-019-0496-8

66. Schneider J, Fischer Fumeaux CJ, Duerden EG, et al. Nutrient Intake in the First Two Weeks of Life and Brain Growth in Preterm Neonates. Pediatrics. 2018;141(3). doi:10.1542/peds.2017-2169

67. Coviello C, Keunen K, Kersbergen KJ, et al. Effects of early nutrition and growth on brain volumes, white matter microstructure, and neurodevelopmental outcome in preterm newborns. Pediatr Res. 2018;83(1-1):102–110. doi:10.1038/pr.2017.227

68. Ottolini KM, Andescavage N, Keller S, Limperopoulos C. Nutrition and the developing brain: the road to optimizing early neurodevelopment: a systematic review. Pediatr Res. 2020;87(2):194–201. doi:10.1038/s41390-019-0508-3

## Supplementary References

1. Wolke D, Ratschinski G, Ohrt B, Riegel K. The cognitive outcome of very preterm infants may be poorer than often reported: an empirical investigation of how methodological issues make a big difference. Eur J Pediatr. 1994;153(12):906–915. doi:10.1007/BF01954744

2. Wolke D, Meyer R. Cognitive status, language attainment, and prereading skills of 6-year-old very preterm children and their peers: the Bavarian Longitudinal Study. Dev Med Child Neurol. 1999;41(2):94–109. doi:10.1017/s0012162299000201

3. Schmitz-Koep B, Zimmermann J, Menegaux A, et al. Decreased amygdala volume in adults after premature birth. Sci Rep. 2021;11(1):5403. doi:10.1038/s41598-021-84906-2

4. Dubowitz LM, Dubowitz V, Goldberg C. Clinical assessment of gestational age in the newborn infant. J Pediatr. 1970;77(1):1–10. doi:10.1016/s0022-3476(70)80038-5

5. Casaer, P., Eggermont, E. Neonatal clinical neurological assessment. In: Harel S, Anastasiow N, eds. The at-risk infant: Psycho/socio/medical aspects. Baltimore, MD: Brookes; 1985:pp. 197– 220.

6. Gutbrod T, Wolke D, Soehne B, Ohrt B, Riegel K. Effects of gestation and birth weight on the growth and development of very low birthweight small for gestational age infants: a matched group comparison. Arch Dis Child Fetal Neonatal Ed. 2000;82(3):F208–14. doi:10.1136/fn.82.3.f208

7. Wang Y, Chen H-J. Use of Percentiles and Z-Scores in Anthropometry. In: Preedy VR, ed. Hand-book of Anthropometry: Physical Measures of Human Form in Health and Disease. 2012nd ed. New York: Springer; 2012.

8. Wit J-M, Boersma B. Catch-up growth: definition, mechanisms, and models. J Pediatr Endocrinol Metab. 2002;15 Suppl 5:1229–1241.

9. Eves R, Mendonça M, Bartmann P, Wolke D. Small for gestational age-cognitive performance from infancy to adulthood: an observational study. BJOG. 2020;127(13):1598–1606. doi:10.1111/1471-0528.16341

10. Voigt M, Fusch C, Olbertz D, et al. Analyse des Neugeborenenkollektivs der Bundesrepublik Deutschland. Geburtsh Frauenheilk. 2006;66(10):956–970. doi:10.1055/s-2006-924458

11. Kromeyer-Hauschild K, Wabitsch M, Kunze D, et al. Perzentile für den Body-mass-Index für das Kindes- und Jugendalter unter Heranziehung verschiedener deutscher Stichproben. Monatsschr Kinderheilkd. 2001;149(8):807–818. doi:10.1007/s001120170107

12. Statistisches Bundesamt. Fragen zur Gesundheit - Körpermaße der Bevölkerung - Mikrozensus 2017. Accessed June 22, 2021. https://www.destatis.de/DE/Themen/Gesellschaft-Umwelt/Gesundheit/Gesundheitszustand-Relevantes-Verhalten/Publikationen/Downloads-Gesund-heitszustand/koerpermasse-5239003179004.html

13. Onis M de, Habicht JP. Anthropometric reference data for international use: recommendations from a World Health Organization Expert Committee. Am J Clin Nutr. 1996;64(4):650–658. doi:10.1093/ajcn/64.4.650

14. Louise T, Nauf Bendar AS, Durighel G, Frost G, Bell J. The effect of preterm birth on adiposity and metabolic pathways and the implications for later life. Clinical Lipidology. 2012;7(3):275–288. doi:10.2217/clp.12.32

15. Euser AM, Wit CC de, Finken MJJ, Rijken M, Wit JM. Growth of preterm born children. Horm Res. 2008;70(6):319–328. doi:10.1159/000161862

16. Olbertz DM, Mumm R, Wittwer-Backofen U, et al. Identification of growth patterns of preterm and small-for-gestational age children from birth to 4 years - do they catch up? J Perinat Med. 2019;47(4):448–454. doi:10.1515/jpm-2018-0239

17. Toftlund LH, Halken S, Agertoft L, Zachariassen G. Catch-Up Growth, Rapid Weight Growth, and Continuous Growth from Birth to 6 Years of Age in Very-Preterm-Born Children. Neonatology. 2018;114(4):285–293. doi:10.1159/000489675

18. Fewtrell MS, Doherty C, Cole TJ, Stafford M, Hales CN, Lucas A. Effects of size at birth, gestational age and early growth in preterm infants on glucose and insulin concentrations at 9-12 years. Diabetologia. 2000;43(6):714–717. doi:10.1007/s001250051368

19. Lapillonne A, Griffin IJ. Feeding preterm infants today for later metabolic and cardiovascular outcomes. J Pediatr. 2013;162(3 Suppl):S7–16. doi:10.1016/j.jpeds.2012.11.048

20. Jain AK. Data clustering: 50 years beyond K-means. Pattern Recognition Letters. 2010;31(8):651–666. doi:10.1016/j.patrec.2009.09.011

21. Likas A, Vlassis N, J. Verbeek J. The global k-means clustering algorithm. Pattern Recognition. 2003;36(2):451–461. doi:10.1016/S0031-3203(02)00060-2

22. Lloyd S. Least squares quantization in PCM. IEEE Transactions on Information Theory. 1982;28(2):129–137. doi:10.1109/TIT.1982.1056489

23. Hedderich DM, Avram M, Menegaux A, et al. Hippocampal subfield volumes are nonspecifically reduced in premature-born adults. Hum Brain Mapp. 2020;41(18):5215–5227. doi:10.1002/hbm.25187

24. Billot B, Bocchetta M, Todd E, Dalca AV, Rohrer JD, Iglesias JE. Automated segmentation of the hypothalamus and associated subunits in brain MRI. Neuroimage. 2020;223:117287. doi:10.1016/j.neuroimage.2020.117287

25. Ronneberger O, Fischer P, Brox T. U-Net: Convolutional Networks for Biomedical Image Segmentation. In: Navab N, Hornegger J, Wells WM, Frangi AF, eds. Medical image computing and computer-assisted intervention - MICCAI 2015: 18th International Conference, Munich, Germany, October 5-9, 2015, proceedings / Nassir Navab, Joachim Hornegger, William M. Wells, Alejandro F. Frangi (eds.). Springer; 2015:234–241.

26. Rodrigues L, Rezende T, Zanesco A, Hernandez AL, Franca M, Rittner L. Hypothalamus fully automatic segmentation from MR images using a U-Net based architecture. In: Romero E, Lepore N, Brieva J, eds. 15th International Symposium on Medical Information Processing and Analysis: 6-8 November 2019, Medellín, Colombia. SPIE; 2020:40.

27. Hayes AF. Introduction to Mediation, Moderation, and Conditional Process Analysis: A Regression-Based Approach. Second edition. New York, NY: The Guilford Press; 2018.

28. Altman DG, Bland JM. How to obtain the P value from a confidence interval. BMJ. 2011;343:d2304. doi:10.1136/bmj.d2304

29. Benjamini Y, Hochberg Y. Controlling the False Discovery Rate: A Practical and Powerful Approach to Multiple Testing. Journal of the Royal Statistical Society. Series B (Methodological). 1995;57(1):289–300. http://www.jstor.org/stable/2346101

30. Bocchetta M, Gordon E, Manning E, et al. Detailed volumetric analysis of the hypothalamus in behavioral variant frontotemporal dementia. J Neurol. 2015;262(12):2635–2642. doi:10.1007/s00415-015-7885-2

31. Neudorfer C, Germann J, Elias GJB, Gramer R, Boutet A, Lozano AM. A high-resolution in vivo magnetic resonance imaging atlas of the human hypothalamic region. Sci Data. 2020;7(1):305. doi:10.1038/s41597-020-00644-6

32. Makris N, Swaab DF, van der Kouwe A, et al. Volumetric parcellation methodology of the human hypothalamus in neuroimaging: normative data and sex differences. Neuroimage. 2013;69:1–10. doi:10.1016/j.neuroimage.2012.12.008

33. Spindler M, Özyurt J, Thiel CM. Automated diffusion-based parcellation of the hypothalamus reveals subunit-specific associations with obesity. Sci Rep. 2020;10(1):22238. doi:10.1038/s41598-020-79289-9

34. Schindler S, Schönknecht P, Schmidt L, et al. Development and evaluation of an algorithm for the computer-assisted segmentation of the human hypothalamus on 7-Tesla magnetic resonance images. PLoS One. 2013;8(7):e66394. doi:10.1371/journal.pone.0066394

35. Gabery S, Georgiou-Karistianis N, Lundh SH, et al. Volumetric analysis of the hypothalamus in Huntington Disease using 3T MRI: the IMAGE-HD Study. PLoS One. 2015;10(2):e0117593. doi:10.1371/journal.pone.0117593

